# Differential DNA Methylation in the Brain as Potential Mediator of the Association between Traffic-related PM_2.5_ and Neuropathology Markers of Alzheimer’s Disease

**DOI:** 10.1101/2023.06.30.23292085

**Authors:** Zhenjiang Li, Donghai Liang, Stefanie Ebelt, Marla Gearing, Michael S. Kobor, Chaini Konwar, Julie L. Maclsaac, Kristy Dever, Aliza Wingo, Allan Levey, James J. Lah, Thomas Wingo, Anke Huels

**Affiliations:** Gangarosa Department of Environmental Health, Rollins School of Public Health, Emory University, 1518 Clifton Rd, Atlanta, GA 30322, USA; Department of Epidemiology, Rollins School of Public Health, Emory University, 1518 Clifton Rd, Atlanta, GA 30322, USA; Department of Pathology and Laboratory Medicine, Emory University, 1364 Clifton Rd, Atlanta, GA 30322, USA; Department of Neurology, Emory University School of Medicine, 12 Executive Park Dr NE, Atlanta, GA 30322, USA; Department of Medical Genetics, University of British Columbia, 4500 Oak St, Vancouver, BC V6H 3N1, Canada; BC Children’s Hospital Research Institute, 938 W 28th Ave, Vancouver, BC V5Z 4H4, Canada; Centre for Molecular Medicine and Therapeutics, 950 W 28th Ave, Vancouver, BC V6H 0B3, Canada; Division of Mental Health, Atlanta VA Medical Center, 1670 Clairmont Rd, Decatur, GA 30033, USA; Department of Psychiatry, Emory University School of Medicine, 12 Executive Park Dr NE #200, Atlanta, GA 30329, USA; Department of Human Genetics, Emory University, 615 Michael Street Suite 301, Atlanta, GA 30322, USA

**Keywords:** Traffic-related fine particulate matter, Alzheimer’s disease, neuropathology, DNA methylation.

## Abstract

**INTRODUCTION:** Growing evidence indicates fine particulate matter (PM_2.5_) as risk factor for Alzheimer’s’ disease (AD), but the underlying mechanisms have been insufficiently investigated. We hypothesized differential DNA methylation (DNAm) in brain tissue as potential mediator of this association.

**METHODS:** We assessed genome-wide DNAm (Illumina EPIC BeadChips) in prefrontal cortex tissue and three AD-related neuropathological markers (Braak stage, CERAD, ABC score) for 159 donors, and estimated donors’ residential traffic-related PM_2.5_ exposure 1, 3 and 5 years prior to death. We used a combination of the Meet-in-the-Middle approach, high-dimensional mediation analysis, and causal mediation analysis to identify potential mediating CpGs.

**RESULTS:** PM_2.5_ was significantly associated with differential DNAm at cg25433380 and cg10495669. Twenty-six CpG sites were identified as mediators of the association between PM_2.5_ exposure and neuropathology markers, several located in genes related to neuroinflammation.

**DISCUSSION:** Our findings suggest differential DNAm related to neuroinflammation mediates the association between traffic-related PM_2.5_ and AD.

## 1 Background

Exposure to traffic-related air pollution (TRAP) is a significant contributor to public health burden with various detrimental health effects.^1^ Fine particulate matter (PM_2.5_), which has been regulated by the National Ambient Air Quality Standards (NAAQS) as a criteria air pollutant since 1997 in the United States (U.S.),^2^ is an important component of TRAP mainly resulting from tailpipe exhaust, brake wear, tire wear, and resuspended dust.^3^ PM_2.5_ from traffic emissions has higher toxicity compared to other natural sources in terms of oxidative potential, cell viability, genotoxicity, oxidative stress, and inflammatory response.^4^ The literature to date demonstrates that exposure to PM_2.5_ is associated with a series of neurological disorders, including dementia and Alzheimer’s disease (AD).^5, 6^

AD is the most common cause of dementia and its hallmark pathologies include accumulation of beta-amyloid (Aβ plaques) outside neurons and aggregation of hyperphosphorylated tau protein (neurofibrillary tangle, NFT) inside neurons in the brain.^7^ In the U.S., 9.30 and 75.68 million people are estimated to develop clinical AD or preclinical AD by 2060,^8^ and the total direct medical costs of AD is estimated to reach $259 billion by 2040.^9^ Due to the growing public concern with these substantial increases in the prevalence of AD, investigations on interventions to prevent progression and onset of AD have targeted the potentially modifiable risk factors of AD, including PM_2.5_.^10^

PM_2.5_ exposure might directly infiltrate the brain^11^ and accelerate AD pathogenesis and development via neuroinflammation, oxidative stress, and Aβ accumulation.^12^ Increasing evidence from human and animal studies proposes that perturbations in DNA methylation (DNAm), which regulates the expression of genes, are associated with indicators of AD as well as PM_2.5_ exposure. However, the tissue specificity of DNAm has limited the ability of previous studies to formally investigate mediation. While there is no conclusive evidence of an association between AD and DNAm in blood,^13^ DNAm alterations in a number of genes were observed to be associated with AD pathology and neuroinflammation in brain tissues, such as amyloid precursor protein (*APP*),^14^ microtubule-associated protein tau (*MAPT*),^14^ apolipoprotein (*APOE*) promoter region,^15^ homeobox A3 (*HOXA3*),^16^ interleukin-1 beta (*IL-1*β),^17^ and interleukin-6 (*IL-6*).^17^

The association of PM_2.5_ with DNAm in blood has been extensively studied.^18^ DNAm in interleukin-10 (*IL-10*), *IL-6*, tumor necrosis factor (*TNF*), toll like receptor 2 (*TLR2*) genes, which play key roles in neuroinflammation,^19^ was reported to be significantly altered in response to short-term exposure to PM_2.5_ and its species.^20^ However, to the best of our knowledge, no human studies have been published on the association between PM_2.5_ exposure and DNAm in human brain, which is the most relevant tissue when studying AD. The only evidence to date comes from *in-vivo* and *in-vitro* studies. Tachibana et al. demonstrated with a mouse model that prenatal exposure to diesel exhaust altered DNAm in brain tissues collected from 1-and 21-day-old offspring, and the differentially methylated CpG sites were enriched in the gene ontology (GO) terms related to neuronal development.^21^ Wei et al. exposed human neuroblastoma cells to near-road PM_2.5_ and found that DNAm was hypermethylated in the promoter regions of genes encoding synaptic neuronal adhesion molecules that mediate essential signaling at synapse.^22^

The mediating role of DNAm for the association between PM_2.5_ and AD pathology has not been well studied, given the limited evidence of an association between PM_2.5_ exposure and DNAm in the brain. The current study investigated the relationship among PM_2.5_, DNAm and AD neuropathology in the post-mortem human brain among brain donors of the Emory Goizueta AD Research Center (ADRC) brain bank. We recently showed a significant association between traffic-related PM_2.5_ exposure and increased AD neuropathology in this dataset.^23^ To elucidate the biological mechanisms for this association, we here investigated whether differential DNAm in the prefrontal cortex tissues mediates the association between long-term exposure to traffic-related PM_2.5_ and the levels of AD-related neuropathological markers. This hypothesis was tested using a combination of the Meet-in-the-Middle (MITM) approach and high-dimensional mediation analysis.

### 2 Methods

#### 2.1 Study design

The current cross-sectional analysis included study participants recruited by the Emory Goizueta ADRC. The ADRC was founded in 2005 and has maintained a brain bank to facilitate AD research. The study participants were research participants evaluated annually, and others were patients treated by Emory Department of Neurology physicians and diagnosed clinically with AD (biomarker defined) or probable AD. The prefrontal cortex tissues were obtained from the participants who had consented to donate biospecimens to the ADRC brain bank. There were 1011 donors enrolled by the third quarter of 2020. After applying the following inclusion criteria, 264 donors remained eligible for the current study: 1) the availability of residential addresses within Georgia (GA) state; 2) age at death equal to or over 55 years (death earlier than 55 possibly due to competing risks); 3) deceased after 1999 (due to the availability of air quality data); 4) no missing values in neuropathology outcomes and key covariates including age at death, race, sex, educational attainment, and APOE genotype. Among these donors, genome-wide DNAm was measured in 161 available samples from the donors deceased after 2007, and after quality control, 159 were included in the current analysis. Written informed consent was provided for all donors, and samples were obtained following research protocols approved by the Emory University Institutional Review Board.

#### 2.2 Neuropathology assessment

The ADRC performed thorough neuropathologic evaluations on the brains of all donors using established comprehensive research evaluations and diagnostic criteria.^24^ These neuropathological assessments include a variety of stains and immunohistochemical preparations, as well as semi-quantitative scoring of multiple neuropathologic changes by experienced neuropathologists using published criteria.^25^ In this project, AD-related neuropathological changes were evaluated using Braak stage, Consortium to Establish a Registry for AD (CERAD) score, and a combination of Amyloid, Braak stage, and CERAD (ABC) score which were developed based on the Aβ plaques and NFTs.^26^ Braak stage is a staging scheme describing NFTs with six stages (Stage I-VI) with a higher stage indicating a wider distribution of NFTs in brain. CERAD score describes the prevalence of Aβ plaques with four levels from no neuritic plaques to frequent. ABC score combines the former two (along with the Thal score for Aβ plaque distribution across various brain regions)^27^ and is transformed into one of four levels: not, low, intermediate, or high level of AD neuropathologic changes.

#### 2.3 Air pollution assessment

Annual concentrations of traffic-related PM_2.5_ were estimated for the 20-county area of Metropolitan Atlanta, GA for 2002-2019. The spatial resolution of the PM_2.5_ data were 250×250m (for 2002-2011) and 200×200m (for 2012-2019). The grid cells of the corresponding side length were evenly distributed throughout the study area. The process for estimating 2002-2011 PM_2.5_ concentrations was previously published.^28, 29^ Briefly, a calibrated Research LINE-source dispersion (R-LINE) model for near surface releases was applied for calculating annual averages of traffic-related PM_2.5_. The model yielded a normalized root mean square error of 24% and a normalized mean bias of 0.3% by comparing with the estimates of the receptor-based source apportionment Chemical Mass Balance Method with Gas Constraints.^28^ For estimating 2012 to 2019 PM_2.5_ concentrations, we trained a land-use random forest model based on the 2015 annual concentrations of traffic-related PM_2.5_ obtained from Atlanta Regional Commission,^30^ road inventory and traffic monitoring data shared by the Georgia Department of Transportation, land cover data accessed via the National Land Cover Database, and ambient PM_2.5_ data obtained from Atmospheric Composition Analysis.^31^ The random forest model was trained with the R package *randomForest*^32^, and two user-defined parameters (i.e., the number of trees and the number of variables randomly tried at each split) were determined by a balance of the efficiency and the out-of-bag R^2^ value. The final model reached an out-of-bag R^2^ of 0.8 and a root-mean-square deviation of 0.2 µg/m^3^. This model was used to predict annual traffic-related PM_2.5_ for 2012-2019 with a spatial resolution of 200m. More details can be found elsewhere.^23^ Finally, we spatially matched geocoded residential addresses to the centroid of closest grids and calculated the individual long-term exposures as the average of specific exposure windows (1 year, 3 years, and 5 years prior to death).

#### 2.4 Genome-wide DNA methylation

DNA was isolated from fresh frozen prefrontal cortex in 161 samples using the QIAGEN GenePure kit. DNAm was assessed with the Illumina Infinium MethylationEPIC BeadChips in batches of 167 prefrontal cortex samples including 6 replicates. The raw intensity files were transformed into a dataset that included beta values for each the CpG sites, and these beta values were computed as the ratio of the methylated signal to the sum of the methylated and unmethylated signals, which ranged from 0 to 1 on a continuous scale. Pre-processing and statistics were done using R (v4.2.0). We followed a validated quality control and normalization pipeline as previously published.^33^ The detailed data processing and sample quality control can be found in the Supplementary Methods. One hundred and fifty-nine samples passed the quality check, and after excluding SNP probes, XY probes and other low-quality probes, 789,286 CpG sites remained. The final DNAm beta values were further normalized to reduce the probe type differences and corrected by *ComBat* to remove the batch effect before the downstream analysis.^34^ We estimated the cell-type proportions (neuronal vs. non-neuronal cells) for each sample using the most recent prefrontal cortex database and the R package *minfi*.^35, 36^

#### 2.5 Covariate assessment

The confounding structure was determined according to literature review and our previous studies, which was illustrated by directed acyclic graphs (DAGs) in the Supplement (Figure S1). Individual-level demographic characteristics [sex, race (Black vs. White), educational attainment (high school or less, college degree, and graduate degree), age at death, APOE ε4 genotype] were obtained from the medical records. APOE ε4 genotype was continuous with a 3-point scale (0 = no ε allele, 1 = one ε4 allele, and 2 = two ε4 alleles). Area Deprivation Index (ADI) for each donor was estimated at the residential address as a proxy for neighborhood socioeconomic status, based on a publicly available database at the level of the Census Block Group for 2015.^37^ Post-mortem interval (hours) of sample collection was provided by our lab collaborators.

#### 2.6 Statistical analysis

Previously, we found higher residential PM_2.5_ exposure was associated with increased AD neuropathology in the Emory Goizueta ADRC brain bank.^23^ To identify DNAm patterns in brain tissue that potentially mediate the association between PM_2.5_ exposure and increased neuropathology markers, we 1) conducted an epigenome-wide association study (EWAS) for the long-term PM_2.5_ exposures 1 year, 3 years, and 5 years prior to death and then investigated whether any differentially methylated CpG sites that were significantly associated with PM_2.5_ exposure in the EWAS were also associated with increased neuropathology markers; and 2) conducted a combination of Meet-in-the-Middle (MITM) approach and high-dimensional mediation analysis (HDMA) to identify any mediating CpGs that did not reach genome-wide significance in the EWAS of PM_2.5_. The MITM approach and HDMA work complementarily to maximize the detention of potential mediators.

Firstly, we conducted an EWAS to assess associations of long-term PM_2.5_ exposures 1 year, 3 years, and 5 years prior to death and methylation levels of CpG sites. Specifically, we used robust multiple linear regression models as implemented in the R package *MASS* to identify differentially CpG sites associated with PM_2.5_ exposures.^38^ To account for measured confounding factors, we included sex, race, educational attainment, age at death, PMI, ADI, and proportion of neuronal cells in the model. Potential batch effect and other unwanted variation were further corrected using the R packages *sva*^39^ (estimating surrogate variables included in the EWAS model as covariates) and *Bacon*.^40^ The *sva* was used to obtain surrogate variables to be included in the models. To account for multiple testing, the *Bonferroni* threshold was used for statistical significance (0.05 / 789,286 = 6.33×10^-8^), while no cut-off was applied on the magnitude of DNA methylation difference.^41^

Any CpG sites that were significantly associated with PM_2.5_ exposure were then investigated for their associations with neuropathology markers. These associations were extracted from an EWAS of each neuropathology marker (CERAD, Braak stage, ABC score) with methylation levels of all CpG sites, using robust multiple linear regression models with the neuropathology markers converted to continuous outcomes and DNAm beta values of CpG sites as exposures, adjusting for sex, race, educational attainment, age at death, PMI, APOE genotype, and proportion of neuronal cells. We used *Bacon*^40^ to control for unmeasured confounding and bias due to the minor inflation/deflation indicated by raw *p*-values.

For the MITM, we compared the 1,000 most significant CpGs from the two sets of EWAS on all CpG sites for PM_2.5_ exposures and neuropathology markers to identify the differentially methylated CpG sties that were associated with both exposures and outcomes. In other words, the raw *p*-values of all 789,286 CpG sites were sorted increasingly, which were derived from the two set of EWAS models conducted on PM_2.5_ exposure and neuropathology markers, respectively. We selected the CpG sites among the lowest 1000 for both PM_2.5_ exposure and neuropathology markers. The MITM approach is widely used in high-dimensional setting to identify intermediate biomarkers.^42^

Then, we conducted a HDMA using the R packages *HIMA* and *DACT* to identify any potential mediating CpG sites between PM_2.5_ exposure and neuropathology from all 789,286 CpG sites. *HIMA* is an R package for estimating and testing high-dimensional mediation effects for omics data, which adopts the multiple mediator model’s framework with reducing the dimensionality of omics data via sure independence screening and minimax concave penalty.^43^ The divide-aggregate composite null test (*DACT*) is a more recent method for HDMA, which utilizes the Efron empirical null framework to calculate a weighted sum of *p*-values obtained from exposure-mediator (EWAS of PM_2.5_ exposure as described above) and mediator-outcome (EWAS of neuropathology markers as described above) models for testing the significance of all mediators^44^. We corrected for multiple testing in *HIMA* and *DACT* using the *Bonferroni* method. Lastly, for the mediating CpG sites identified by either *HIMA* or *DACT*, we used the R package *mediation* to conduct a causal mediation analysis obtain their indirect effects.^45–47^ The *mediation* is a frequently used tool which implements the mediation methods and suggestions proposed by Imai et al.^48, 49^ The average causal mediation effect (i.e., indirect effect) and total effect estimated by *mediation* were summarized for the CpG sites with positive indirect effects that were in line with the hypothesized adverse effect of traffic-related PM_2.5_ on neuropathology markers. In contrast to the MITM approach described earlier, HDMA examine multiple mediators together in a framework of mediation analysis, which allowed us to ascertain the extent to which the particular indirect effects were associated with the mediators.

To aid the interpretation of model results, we conducted a gene ontology analysis using the R package *missMethyl* based on the top 1000 CpG sites with lowest raw *p*-values^50^. The gene ontology analysis was conducted for the EWAS results of PM_2.5_ exposure as well as for the EWAS results of the three neuropathology markers. All CpG sites were annotated using an online annotation data for the ‘IlluminaHumanMethylationEPIC’.^51^ Additional functional insight on single CpG sites was obtained by searching the corresponding CpG site in publicly available databases, including EWAS catalog^52^.

All analyses were completed in R (v4.2.0).

### 3 Results

#### 3.1 Study population characteristics

A total of 159 donors were included in the current analysis, and their demographic characteristics and neuropathologic markers are described in Table 1. The average age of death was 76.6 years (SD=9.98) and 56% of the study population were male. The study population was predominantly white (89.3%) and well-educated with 123 (78.7%) completing college or more and living in less deprived neighborhoods (ADI: mean = 36.3, SD = 24.2). The majority of study sample (95.6%) were diagnosed with AD or other forms of dementia, and the prevalence of the APOE ε4 allele (56% with at least one APOE ε4 allele) in this population was much higher than that in the general population in the U.S.^53^

**Table 1.**
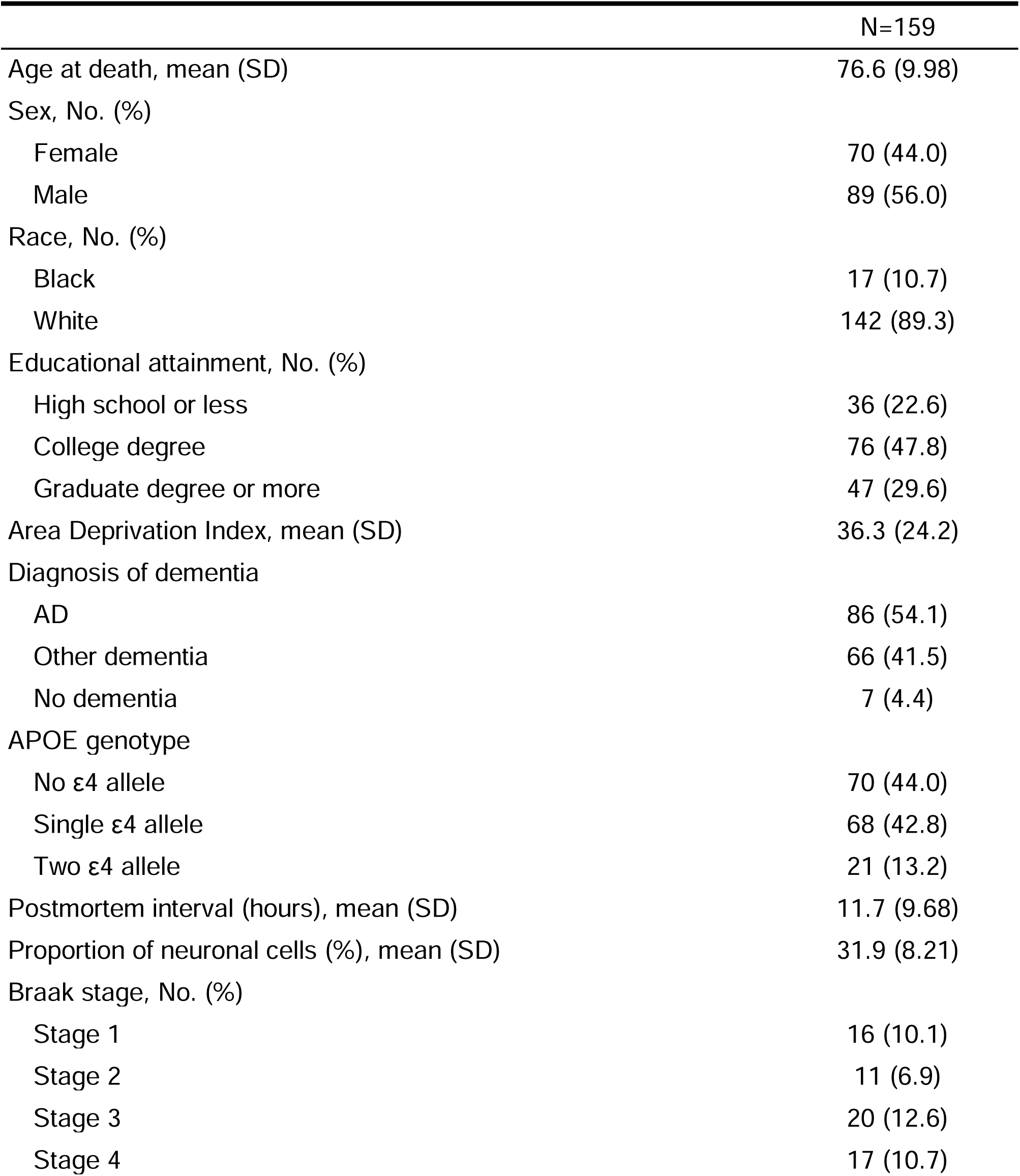
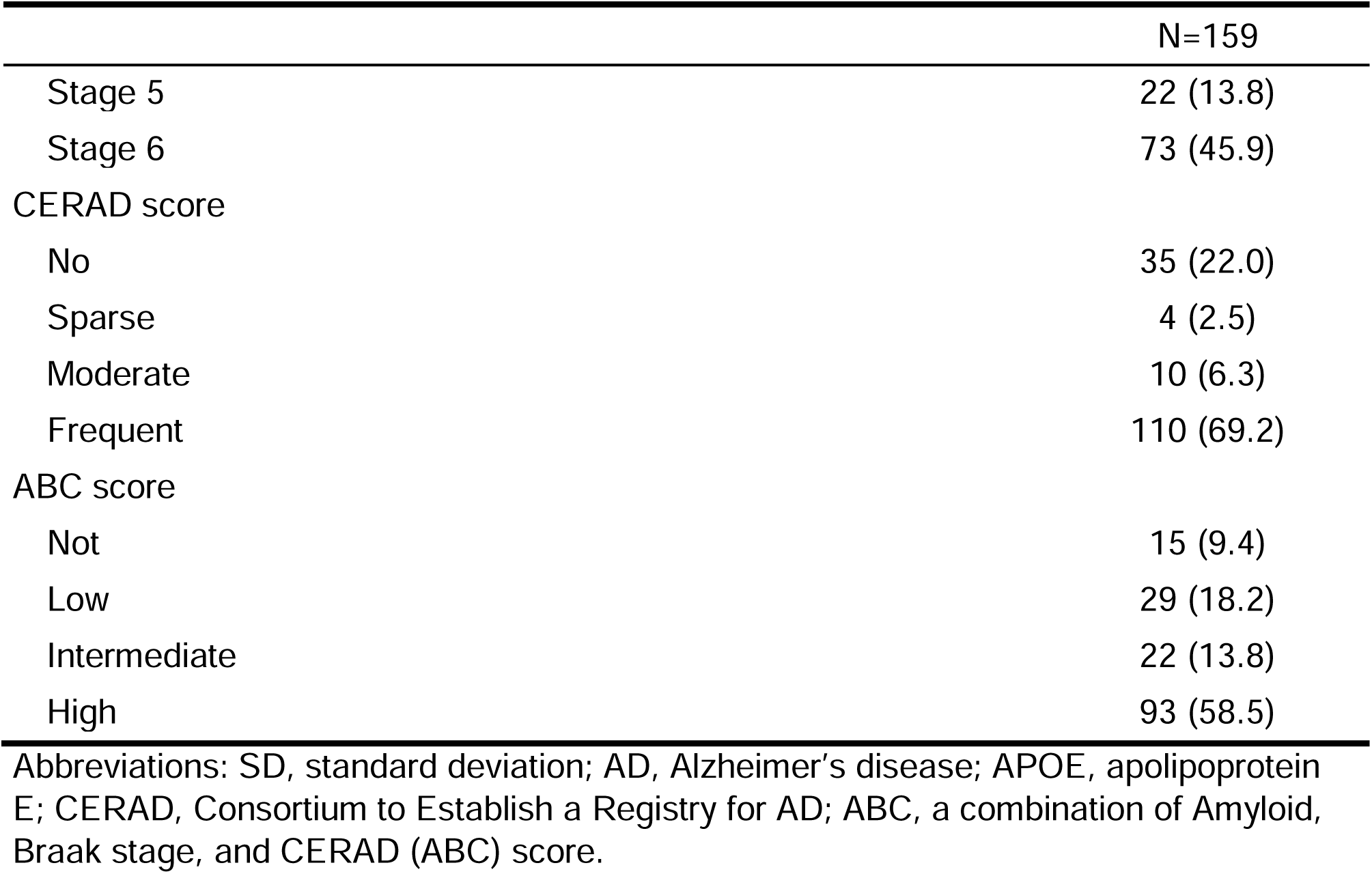
Selected population characteristics among the donors included in the current analysis.

|  | N=159 |
| --- | --- |
| Age at death, mean (SD) | 76.6 (9.98) |
| Sex, No. (%) |  |
| Female | 70 (44.0) |
| Male | 89 (56.0) |
| Race, No. (%) |  |
| Black | 17 (10.7) |
| White | 142 (89.3) |
| Educational attainment, No. (%) |  |
| High school or less | 36 (22.6) |
| College degree | 76 (47.8) |
| Graduate degree or more | 47 (29.6) |
| Area Deprivation Index, mean (SD) | 36.3 (24.2) |
| Diagnosis of dementia |  |
| AD | 86 (54.1) |
| Other dementia | 66 (41.5) |
| No dementia | 7 (4.4) |
| APOE genotype |  |
| No ε4 allele | 70 (44.0) |
| Single ε4 allele | 68 (42.8) |
| Two ε4 allele | 21 (13.2) |
| Postmortem interval (hours), mean (SD) | 11.7 (9.68) |
| Proportion of neuronal cells (%), mean (SD) | 31.9 (8.21) |
| Braak stage, No. (%) |  |
| Stage 1 | 16 (10.1) |
| Stage 2 | 11 (6.9) |
| Stage 3 | 20 (12.6) |
| Stage 4 | 17 (10.7) |
| Stage 5 | 22 (13.8) |
| Stage 6 | 73 (45.9) |
| CERAD score |  |
| No | 35 (22.0) |
| Sparse | 4 (2.5) |
| Moderate | 10 (6.3) |
| Frequent | 110 (69.2) |
| ABC score |  |
| Not | 15 (9.4) |
| Low | 29 (18.2) |
| Intermediate | 22 (13.8) |
| High | 93 (58.5) |

Abbreviations: SD, standard deviation; AD, Alzheimer's disease; APOE, apolipoprotein E; CERAD, Consortium to Establish a Registry for AD; ABC, a combination of Amyloid, Braak stage, and CERAD (ABC) score.

As illustrated by the 1-year traffic-related PM_2.5_ exposure (Figure 1A), donors living in urban areas had a higher level of PM_2.5_ exposure compared to those living in suburban areas. The median of 1-year exposure was 1.21 µg/m^3^ [interquartile range (IQR)=0.78]. As PM_2.5_ concentrations have decreased over the last decades, 3-year and 5-year exposures were slightly higher (3-year exposure: median=1.32 µg/m^3^ [IQR=0.74], 5-year exposure: median=1.39 µg/m^3^ [IQR: 0.81]) (Figure 1B).

**Figure 1.**
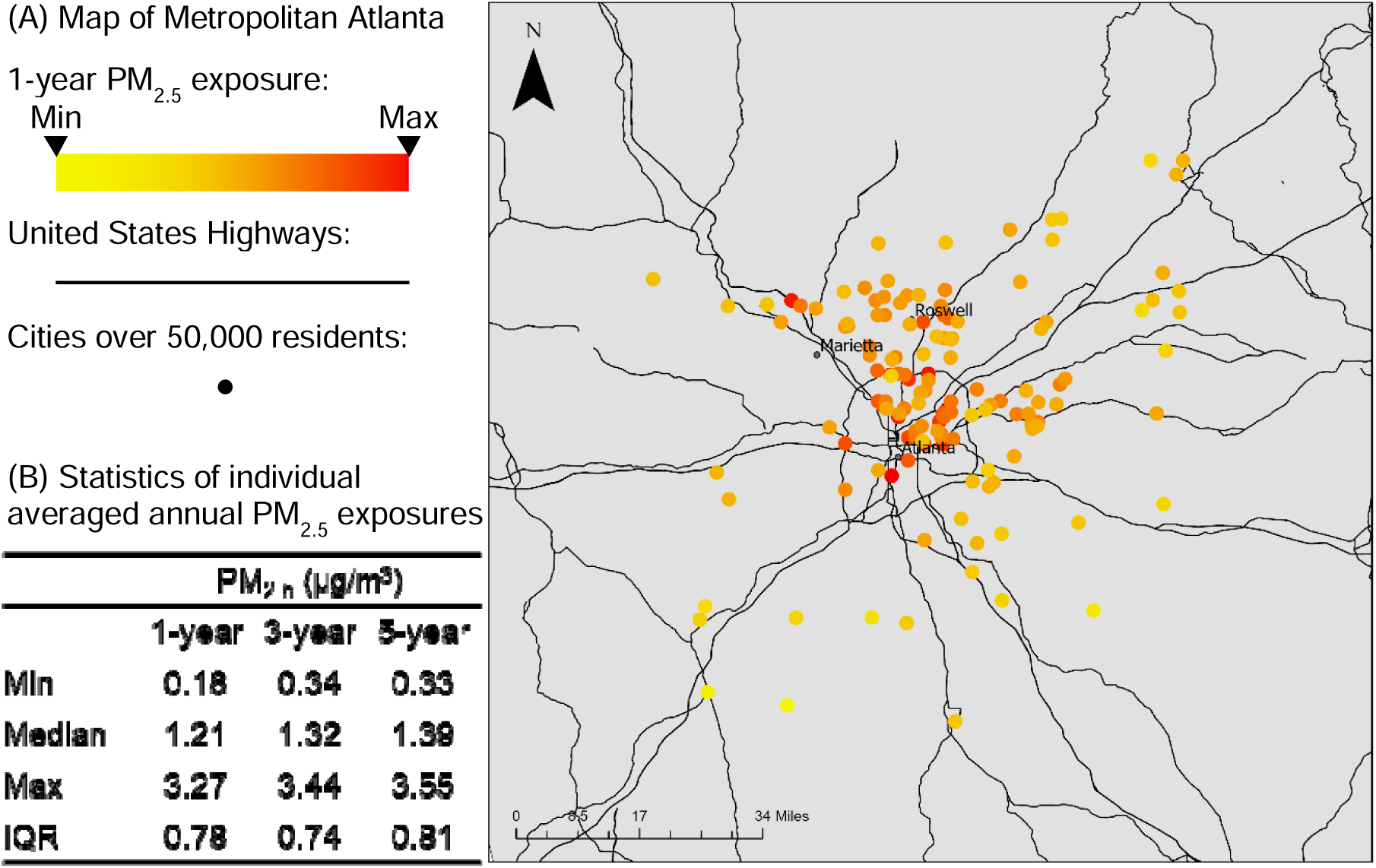
Statistics and distribution of PM_2.5_ exposures in Metropolitan Atlanta (study area), Georgia, United States. (A) Map of Metropolitan Atlanta with individual 1-year averaged annual PM_2.5_ exposure. The dots denote the donors’ residential address and are colored according to their PM_2.5_ exposures as showed in the legend. Red means a higher exposure level. (B) Statistics of individual averaged annual PM_2.5_ exposures for 1 year, 3 years, and 5 years.

#### 3.2 Association between PM_2.5_ exposure and DNAm in the brain

After correcting for multiple tests and adjusting for bias and measured and unmeasured confounding, two CpG sites (cg25433380 and cg10495669) were consistently associated with PM_2.5_ across different exposure windows (Figure 2, Table 2; summary statistics for all 789,286 CpG sites are provided as Tables S4-6 in spreadsheets). For example, a 1 µg/m^3^ increase in 1-year PM_2.5_ exposure was associated with 0.0065 increase in the DNAm beta value of cg25433380 (*p* = 1.58×10^-8^). cg25433380 and cg10495669 are on chromosome 9 and 20, respectively, and cg10495669 is assigned to the gene encoding RanBP-type and C3HC4-type zinc finger-containing protein 1 (*RBCK1*). The two CpG sites were not significantly associated with any neuropathology markers (Table 2).

**Figure 2.**
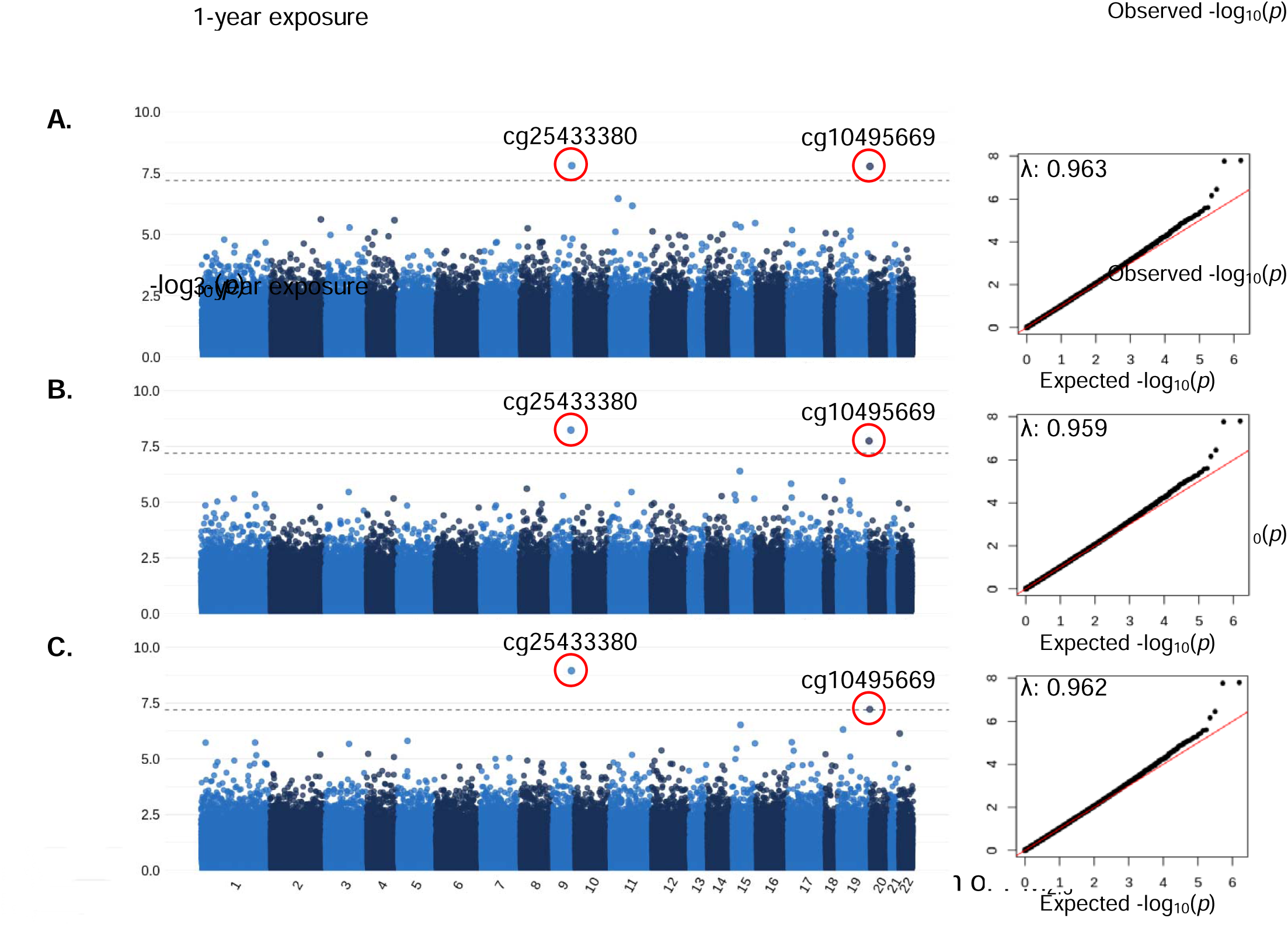
Manhattan and QQ plots for the epigenome-wide association of PM_2.5_ exposures (A. 1-year / B. 3-year / C. 5-year average exposure prior to death) and DNA methylation in postmortem frontal cortex tissue. λ denotes the inflation factor. Adjusted for covariates: age at death, sex, race, educational attainment, post-mortem interval, area deprivation index, and cell type composition. Unmeasured confounding and bias were adjusted with surrogate variable analysis and R package *Bacon*. Bonferroni threshold: 0.05/789,286.

**Table 2.** CpGs associated with traffic-related PM_2.5_ exposure prior to death and their association with neuropathology markers.

| CpG | chr | Position | Gene |  | Coefficients <sup>a</sup> | p-values <sup>b</sup> |
| --- | --- | --- | --- | --- | --- | --- |
| A. CpGs with PM<sub>2.5</sub> exposures |  |  |  |  |  |  |
| cg25433380 | 9 | 388,531 | Intergenic | 1-year exposure | 0.0065 | 1.58×10<sup>-8</sup> |
|  |  |  |  | 3-year exposure | 0.0066 | 5.82×10<sup>-9</sup> |
|  |  |  |  | 5-year exposure | 0.0063 | 1.12×10<sup>-9</sup> |
| cg10495669 | 20 | 137,531,767 | RBCK1 | 1-year exposure | 0.0127 | 1.69×10<sup>-8</sup> |
|  |  |  |  | 3-year exposure | 0.0128 | 1.78×10<sup>-8</sup> |
|  |  |  |  | 5-year exposure | 0.0114 | 5.96×10<sup>-8</sup> |
| B. CpGs with neuropathology markers |  |  |  |  |  |  |
| cg25433380 | 9 | 388,531 | Intergenic | Braak stage | 0.08 | 0.729 |
|  |  |  |  | CERAD | 0.05 | 0.629 |
|  |  |  |  | ABC | 0.04 | 0.825 |
| cg10495669 | 20 | 137,531,767 | RBCK1 | Braak stage | 0.02 | 0.929 |
|  |  |  |  | CERAD | 0.12 | 0.397 |
|  |  |  |  | ABC | 0.09 | 0.593 |

Abbreviations: PM<sub>2.5</sub>, fine particulate matter; chr, chromosome; RBCK1, RanBP-type and C3HC4-type zinc finger-containing protein 1.
<sup>a</sup> The coefficients for PM<sub>2.5</sub> exposures represent the change in the beta values of CpG sites associated with one-unit increase in the exposures; the coefficients for neuropathology markers represent the change in the neuropathology markers associated with one-interquartile-range increase in the beta values of CpG sites.
<sup>b</sup> The Bonferroni threshold: $0.05/789,286 \approx 6.33 \times 10^{-8}$ .

#### 3.3 Meet-in-the-Middle approach and high-dimensional mediation analysis

For the MITM approach, we explored the overlapping CpG sites among the top 1000 CpG sites for the EWAS of PM_2.5_ and the EWAS of neuropathology markers (results presented in Tables S7-S9 in spreadsheets) and identified four overlapping CpG sites (Table S1). Specifically, DNAm in cg01835635 (apolipoprotein A4 gene, *APOA4*) was associated with CERAD score as well as PM_2.5_ exposure for the 1-year and 3-year exposure windows. DNAm in cg09830308 (mixed lineage kinase domain like pseudokinase gene, *MLKL*) was associated with Braak stage as well as PM_2.5_ exposures for the 1-year, 3-year, and 5-year windows; cg16342341 (sorbin and SH3 domain-containing protein 2 gene, *SORBS2*) was associated with CERAD score as well as 1-year PM_2.5_ exposure; and cg27459981 (*MLKL* gene) was associated with Braak stage and ABC score as well as PM_2.5_ exposures for the 3-year and 5-year windows.

The HDMA via *HIMA* did not identify any CpG sites as significant mediators. In the HDMA using a combination of *DACT* and causal mediation analysis, we identified twenty-two CpG sites to mediate the positive association between PM_2.5_ exposure and ABC score (Table 3), while none were observed for Braak stage and CERAD score.

**Table 3.**
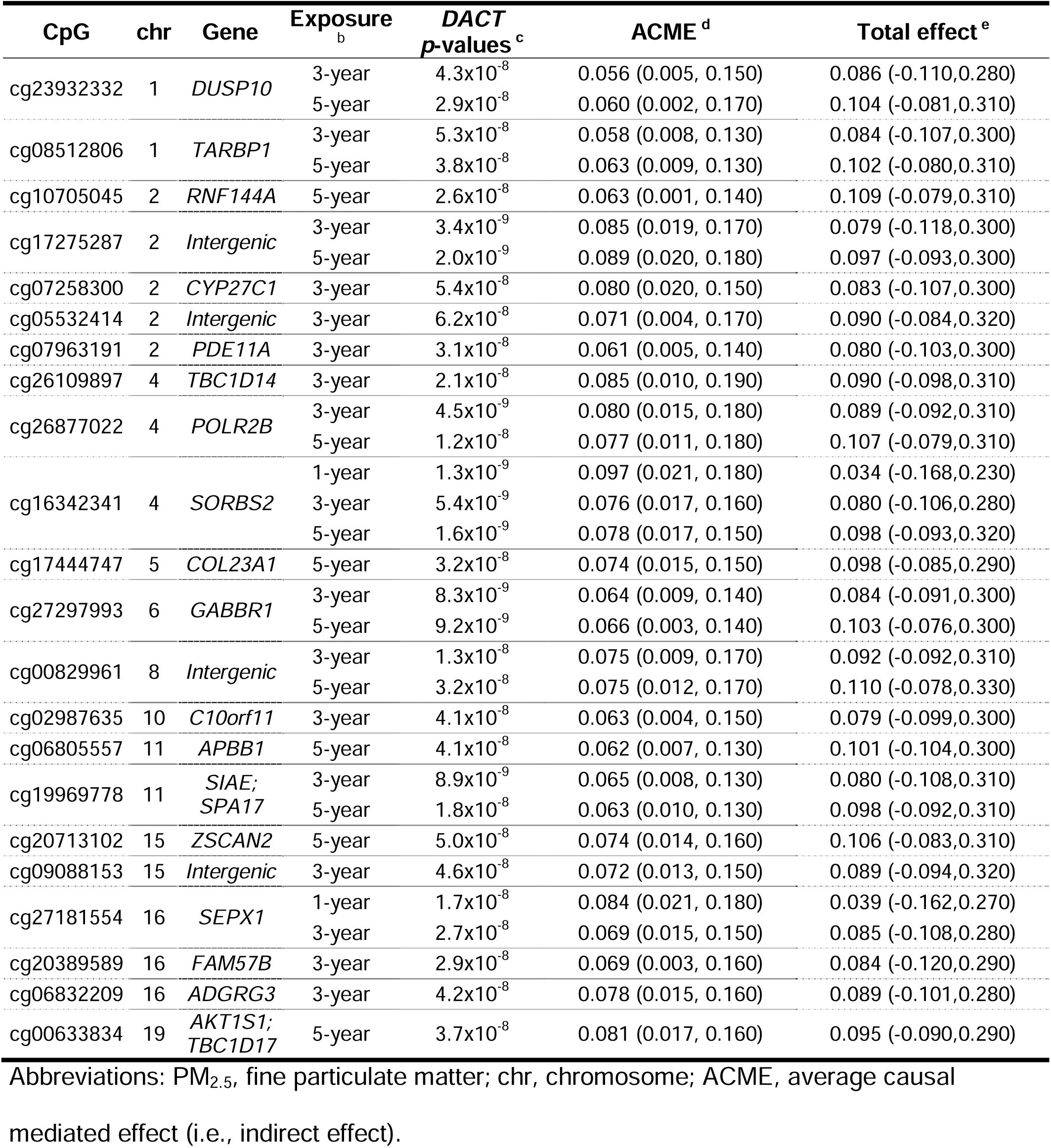
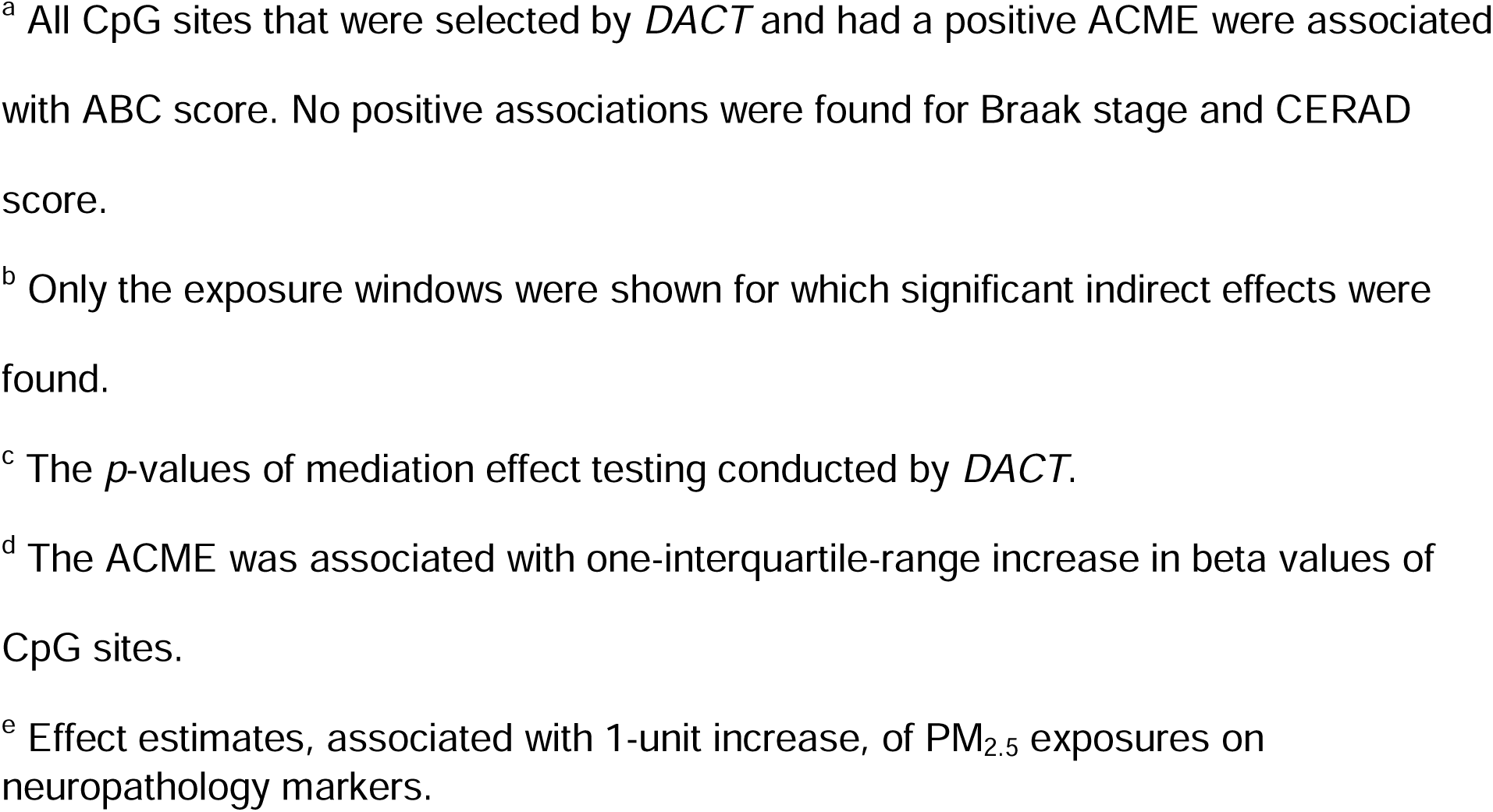
Indirect effect estimated by causal mediation analysis via the R package *mediation* of CpG sites selected by high-dimensional mediation analysis for the associations between PM_2.5_ exposure and ABC score ^a^.

| CpG | chr | Gene | Exposure<sup>b</sup> | DACT<br>p-values<sup>c</sup> | ACME<sup>d</sup> | Total effect<sup>e</sup> |
| --- | --- | --- | --- | --- | --- | --- |
| cg23932332 | 1 | DUSP10 | 3-year | 4.3x10<sup>-8</sup> | 0.056 (0.005, 0.150) | 0.086 (-0.110,0.280) |
|  |  |  | 5-year | 2.9x10<sup>-8</sup> | 0.060 (0.002, 0.170) | 0.104 (-0.081,0.310) |
| cg08512806 | 1 | TARBP1 | 3-year | 5.3x10<sup>-8</sup> | 0.058 (0.008, 0.130) | 0.084 (-0.107,0.300) |
|  |  |  | 5-year | 3.8x10<sup>-8</sup> | 0.063 (0.009, 0.130) | 0.102 (-0.080,0.310) |
| cg10705045 | 2 | RNF144A | 5-year | 2.6x10<sup>-8</sup> | 0.063 (0.001, 0.140) | 0.109 (-0.079,0.310) |
| cg17275287 | 2 | Intergenic | 3-year | 3.4x10<sup>-9</sup> | 0.085 (0.019, 0.170) | 0.079 (-0.118,0.300) |
|  |  |  | 5-year | 2.0x10<sup>-9</sup> | 0.089 (0.020, 0.180) | 0.097 (-0.093,0.300) |
| cg07258300 | 2 | CYP27C1 | 3-year | 5.4x10<sup>-8</sup> | 0.080 (0.020, 0.150) | 0.083 (-0.107,0.300) |
| cg05532414 | 2 | Intergenic | 3-year | 6.2x10<sup>-8</sup> | 0.071 (0.004, 0.170) | 0.090 (-0.084,0.320) |
| cg07963191 | 2 | PDE11A | 3-year | 3.1x10<sup>-8</sup> | 0.061 (0.005, 0.140) | 0.080 (-0.103,0.300) |
| cg26109897 | 4 | TBC1D14 | 3-year | 2.1x10<sup>-8</sup> | 0.085 (0.010, 0.190) | 0.090 (-0.098,0.310) |
| cg26877022 | 4 | POLR2B | 3-year | 4.5x10<sup>-9</sup> | 0.080 (0.015, 0.180) | 0.089 (-0.092,0.310) |
|  |  |  | 5-year | 1.2x10<sup>-8</sup> | 0.077 (0.011, 0.180) | 0.107 (-0.079,0.310) |
| cg16342341 | 4 | SORBS2 | 1-year | 1.3x10<sup>-9</sup> | 0.097 (0.021, 0.180) | 0.034 (-0.168,0.230) |
|  |  |  | 3-year | 5.4x10<sup>-9</sup> | 0.076 (0.017, 0.160) | 0.080 (-0.106,0.280) |
|  |  |  | 5-year | 1.6x10<sup>-9</sup> | 0.078 (0.017, 0.150) | 0.098 (-0.093,0.320) |
| cg17444747 | 5 | COL23A1 | 5-year | 3.2x10<sup>-8</sup> | 0.074 (0.015, 0.150) | 0.098 (-0.085,0.290) |
| cg27297993 | 6 | GABBR1 | 3-year | 8.3x10<sup>-9</sup> | 0.064 (0.009, 0.140) | 0.084 (-0.091,0.300) |
|  |  |  | 5-year | 9.2x10<sup>-9</sup> | 0.066 (0.003, 0.140) | 0.103 (-0.076,0.300) |
| cg00829961 | 8 | Intergenic | 3-year | 1.3x10<sup>-8</sup> | 0.075 (0.009, 0.170) | 0.092 (-0.092,0.310) |
|  |  |  | 5-year | 3.2x10<sup>-8</sup> | 0.075 (0.012, 0.170) | 0.110 (-0.078,0.330) |
| cg02987635 | 10 | C10orf11 | 3-year | 4.1x10<sup>-8</sup> | 0.063 (0.004, 0.150) | 0.079 (-0.099,0.300) |
| cg06805557 | 11 | APBB1 | 5-year | 4.1x10<sup>-8</sup> | 0.062 (0.007, 0.130) | 0.101 (-0.104,0.300) |
| cg19969778 | 11 | SIAE;<br>SPA17 | 3-year | 8.9x10<sup>-9</sup> | 0.065 (0.008, 0.130) | 0.080 (-0.108,0.310) |
|  |  |  | 5-year | 1.8x10<sup>-8</sup> | 0.063 (0.010, 0.130) | 0.098 (-0.092,0.310) |
| cg20713102 | 15 | ZSCAN2 | 5-year | 5.0x10<sup>-8</sup> | 0.074 (0.014, 0.160) | 0.106 (-0.083,0.310) |
| cg09088153 | 15 | Intergenic | 3-year | 4.6x10<sup>-8</sup> | 0.072 (0.013, 0.150) | 0.089 (-0.094,0.320) |
| cg27181554 | 16 | SEPX1 | 1-year | 1.7x10<sup>-8</sup> | 0.084 (0.021, 0.180) | 0.039 (-0.162,0.270) |
|  |  |  | 3-year | 2.7x10<sup>-8</sup> | 0.069 (0.015, 0.150) | 0.085 (-0.108,0.280) |
| cg20389589 | 16 | FAM57B | 3-year | 2.9x10<sup>-8</sup> | 0.069 (0.003, 0.160) | 0.084 (-0.120,0.290) |
| cg06832209 | 16 | ADGRG3 | 3-year | 4.2x10<sup>-8</sup> | 0.078 (0.015, 0.160) | 0.089 (-0.101,0.280) |
| cg00633834 | 19 | AKT1S1;<br>TBC1D17 | 5-year | 3.7x10<sup>-8</sup> | 0.081 (0.017, 0.160) | 0.095 (-0.090,0.290) |

Abbreviations: PM<sub>2.5</sub>, fine particulate matter; chr, chromosome; ACME, average causal mediated effect (i.e., indirect effect).
<sup>a</sup> All CpG sites that were selected by *DACT* and had a positive ACME were associated with ABC score. No positive associations were found for Braak stage and CERAD score.
<sup>b</sup> Only the exposure windows were shown for which significant indirect effects were found.
<sup>c</sup> The *p*-values of mediation effect testing conducted by *DACT*.
<sup>d</sup> The ACME was associated with one-interquartile-range increase in beta values of CpG sites.
<sup>e</sup> Effect estimates, associated with 1-unit increase, of PM<sub>2.5</sub> exposures on neuropathology markers.

One CpG site (cg16342341, *SORBS2* gene) was associated with all three exposure windows (1, 3 and 5-years prior to death), and eight with two exposure windows. Of note, cg16342341 (*SORBS2*) was also identified in the MITM approach described above. The total effect estimated for all mediation analyses was positive but insignificant in this subsample of the cohort (see Christensen et al. 2023 for the significant total effect in the full cohort).^23^ The summary statistics for all CpG sites detected by *DACT* are summarized in the Supplement (Table S2).

#### 3.4 Secondary analyses

A gene ontology analysis was conducted for the top 1000 CpG sites associated PM_2.5_ and for the top 1000 CpG sites associated with the neuropathology markers. None of the KEGG pathways reached significance after correcting for multiple tests. Therefore, we summarized the top 10 KEGG pathways for each of the PM_2.5_ exposures or neuropathology markers in the Supplement (Table S3). One pathway, which is the longevity regulating pathway, was associated with both 3-year exposure to PM_2.5_ and CERAD score. Eight genes (*HSPA1A*, *HSPA1L*, *IRS1*, *KRAS*, *NRAS*, *RPTOR*, *IRS2*, *ATG5*) in this pathway were enriched by differentially methylated CpG sites that were associated with 3-year PM_2.5_ exposure, and ten genes (*ADCY3*, *ADCY5*, *NFKB1*, *PRKAG2*, *RPTOR*, *TSC2*, *EHMT1*, *ULK1*, *AKT1S1*, *ATG5*) with CERAD score. Of note, *AKT1S1* was also among the genes that were identified in the HDMA (DACT and causal mediation analysis).

## 4 Discussion

In the current study of 159 donors from the Emory Goizueta ADRC brain bank, we identified differential DNAm in prefrontal cortex tissues at two CpG sites to be significantly associated with long-term PM_2.5_ exposure. The two CpG sites (cg25433380 and cg10495669) that were associated with PM_2.5_ exposure were consistently associated with long-term exposures to traffic-related PM_2.5_ 1 year, 3 years, and 5 years prior to death, after controlling for measured and unmeasured confounding. While cg25433380 and cg10495669 were not associated with increases in neuropathology markers, we identified 4 CpG sites that overlapped between the top 1000 CpG sites associated with PM_2.5_ and neuropathology markers (MITM approach) and 22 CpG sites that mediated the adverse effect of PM_2.5_ exposures on AD-related neuropathology markers using HDMA. The longevity regulating pathway was enriched by differentially methylated CpG sites associated with PM_2.5_ (3-year exposure window) and CERAD score.

This is the first study showing an association between PM_2.5_ exposure and differential DNAm in the brain (cg25433380 and cg10495669). Scarce evidence related to air pollution has been reported on cg25433380. Higher DNA methylation levels of cg10495669 in nasal cells have been associated with 1-year ambient PM_2.5_ exposure among 503 children in Massachusetts.^54^ *RBCK1*, the gene which cg10495669 is assigned to, is involved in carcinogenesis and inflammation pathways. Yu et al. suggested that *RBCK1* promoted the ubiquitination and degradation of p53.^55^ The impairment of p53 expression and activity might participate in neurodegeneration, as p53 can bind to genes that regulate expression of synaptic proteins, neurite outgrowth, and axonal regeneration, which indicated a neuroprotective role against AD development.^56^ In addition, *RBCK1* can regulate the proinflammatory-cytokines-induced nuclear factor kappa B (NF-kB) activation which serves as a pivotal mediator of inflammatory responses.^57^ NF-kB activation is a common feature of many neurodegenerative diseases,^58^ and the increased expression and/or activation of NF-kB has been largely observed in post-mortem studies of AD patients.^59^ However, the two CpG sites were not found to be associated with any neuropathology markers in the current analysis. More research is warranted on these CpG sites to investigate their potential role in AD development with a larger sample size and participants of more diverse disease stages from preclinical to severe dementia.

We identified four CpG sites (cg01835635, cg09830308, cg16342341, and cg27459981) that overlapped between the top 1000 CpG sites associated with both PM_2.5_ and neuropathology markers via MITM approach. Three of them (cg16342341, cg09830308 and cg27459981) or their related genes have been previously associated with AD or PM_2.5_ exposure. Cg09830308 and cg27459981, assigned to *MLKL*, were both associated with Braak stage and PM_2.5_ exposure 3 and 5 years prior to death. *MLKL* plays a critical role in TNF-induced cell death (i.e., necroptosis). Caccamo et al. found that necroptosis was activated in postmortem brains of AD patients and positively correlated with Braak stage, and *MLKL* expression was significantly higher compared to control cases’ brain tissues.^60^ Similarly, Jayaraman et al. reported that necroptosis signaling was highly activated in the hippocampus of AD patients, as illustrated by the increased mRNA expression of genes, including *MLKL*.^61^ Furthermore, Wang et al. demonstrated that the knockdown of *MLKL* significantly increased the ratio of Aβ42 to Aβ40, a potential diagnostic marker of AD, in an AD model HEK293 cell line.^62^ Collectively, PM_2.5_ exposure might induce the TNF-mediated neuroinflammation, resulting in necroptosis, and thus contribute to AD pathogenesis.

Cg16342341, assigned to *SORBS2*, was also identified as a potential mediator in the HDMA, where it mediated the association of all PM_2.5_ exposure windows with ABC score. As *SORBS2* is well known for its role in AD and neuroinflammation^63, 64^ and was associated with PM_2.5_ exposure in rats^65^, our findings contribute to the evidence of *SORBS2* playing a role in PM_2.5_-induced neuropathologic changes of AD. *SORBS2* represses IL-6 and TNF-α expression in the mouse embryonic fibroblasts,^66^ and Chen et al. demonstrated that its level was lower in the brains of AD model mice compared to wild type mice,^63^ implying a role of *SORBS2* in regulating neuroinflammation. In a human study, genetic variation in *SORBS2* was associated with age at onset of AD.^64^ While evidence on the association between PM_2.5_ exposure and *SORBS2* is more scarce, Chao et al. reported that prenatal exposure to PM_2.5_ induced upregulation of microRNAs targeting *SORBS2* gene in fetal rat cortex tissues.^65^

We also identified 21 other CpGs as potential mediators of the association between long-term exposure to traffic-related PM_2.5_ and ABC score using HDMA, and two of these CpGs (cg07963191 and cg27297993) have been previously reported in association with AD. Cg07963191 was assigned to the dual 3’,5’-cyclic-AMP and -GMP phosphodiesterase 11A gene (*PDE11A*) that participates in neuroplasticity and neuroprotection.^67^ Cg27297993 was assigned to the gamma-aminobutyric acid B receptor 1 gene (*GABBR1*). *GABBR1* is the main inhibitory neurotransmitter, which was reported to be downregulated in the brains of AD patients.^68^ Iwakiri et al. observed a negative correlation between *GABBR1* and NFT formation in the hippocampus of seniors, suggesting an increased or stable expression of *GABBR1* may contribute to neuronal resistance to AD development.^69^

To derive more functional insights, we conducted gene ontology analysis based on KEGG pathway database for the top 1000 CpGs associated with PM_2.5_ exposure or neuropathology markers.^50^ Proline-rich AKT1 substrate 1 (*AKT1S1*) was one of the genes enriched in the longevity regulating pathway, an overlapping pathway between PM_2.5_ exposure and CERAD score. Of note, differential DNA methylation in cg00633834, which is assigned to *AKT1S1*, was also identified in HDMA. *AKT1S1* can activate mammalian target of rapamycin (mTOR)–mediated signaling pathways when phosphorylated,^70^ and mTOR signaling was observed to have higher activity in AD brains, suggesting a role of *AKT1S1* in the accumulation of Aβ and tau proteins.^71^

The current analysis employed the MITM approach and HDMA simultaneously to maximize the potential of identifying the differentially methylated CpG sites lying on a pathway from PM_2.5_ to AD-related neuropathology. The application of the MITM approach was based on the investigation of epigenomics vs. PM_2.5_ exposures and AD-related neuropathology vs. epigenomics, which lent credibility to the association between PM_2.5_ exposure and AD-related neuropathology by breaking it down and linking it up with DNAm.^72^ Furthermore, while conventional methods of multiple testing correction (e.g., Bonferroni method) may overlook potential relevant CpG sites, especially given a small sample size, the MITM approach serves as a supplement by taking into account the biological relevance regardless of their statistical significance.^72^ However, the MITM approach assumes that all intermediate variables are independent, which is not always the case in real-world scenarios. The HDMA focuses more on quantifying the indirect effect of the mediator and considers the potential correlation among mediators.^43^ Admittedly, we did not observe many consistencies, except for cg16342341 (*SORBS2*), between the two approaches.

Our study has several strengths. We established for the first time a potential mediation effect of DNAm for the association between PM_2.5_ and neuropathological changes of AD. The neuropathological changes of AD were quantified via multiple markers, including Braak stage, CERAD score, and ABC score, which covers the essential components (i.e., NFTs and Aβ plaques) for the neuropathological diagnosis of AD. Further, the neuropathology markers were assessed by experienced neuropathologists at Emory Goizueta ADRC following a standardized protocol, which minimized the misclassification bias of outcomes. Finally, the high-resolution PM_2.5_ exposure assessment model enabled the characterization of spatial variation in individual exposure and reduced the potential measurement error.^73^

Our study has several limitations. First, the temporal sequence between DNAm changes and AD neuropathology could not be clearly defined because both were assessed post-mortem. Second, traffic-related PM_2.5_ exposure was estimated based on the residential address of donors at death. Moving shortly prior to death could have introduced measurement errors in exposure assessment. The selection of exposure windows was arbitrary, as the disease process of AD may start many years before death and vary by patients. Third, the results were from a single brain bank and participants with a high APOE ε4 carrier rate, so the generalizability should be tested in other brain banks or autopsy cohorts. Fourth, even though most of the study population was White, and we controlled for race, the ancestry effect on DNA methylation might persist as residual confounding. Fifth, the current analysis only focused on the effects of PM_2.5_, while other air pollutants such as nitrogen oxides or ozone might also play a role for AD.^74, 75^ Finally, while the sample size of 159 brain samples was relatively large considering the challenges in collecting such samples, the high dimensionality of the genome-wide DNAm data raises concerns about the reliability of our findings.

Our findings provide important information on the biological mechanisms underlying the PM_2.5_ toxicity on AD pathogenesis. Future studies evaluating the mediating role of DNAm on AD-related outcomes should consider: 1) performing the analysis among early-stage AD patients or patients with mild cognition impairment to further illustrate the role of PM_2.5_ in AD etiology; 2) performing genome-wide DNAm together with transcriptomics, proteomics, and/or metabolomics to capture a holistic picture of the underlying mechanism.

## Data Availability

All data produced in the present study are available upon reasonable request to the authors.

## Consent Statement

Written informed consent was provided for all subjects, and samples were obtained following research protocols approved by the Emory University Institutional Review Board.

## Acknowledgements

We want to thank Dr. Jeremy A. Sarnat (Emory), Dr. Armistead Russell (Georgia Tech), Ms. Kyung-Hwa Kim and Ms. Abby Marinelli from the Atlanta Regional Commission for providing data and guidance towards the air pollution exposure assessment.

## References

1. Fox M, Koehler K, Johnson N. Established and emerging effects of traffic-related air pollution. Traffic-Related Air Pollution. Elsevier; 2020:207–228.

2. Bachmann J. Will the circle be unbroken: a history of the U.S. National Ambient Air Quality Standards. J Air Waste Manag Assoc. Jun 2007;57(6):652–97. doi:10.3155/1047-3289.57.6.652

3. Jeong CH, Wang JM, Hilker N, et al. Temporal and spatial variability of traffic-related PM2.5 sources: Comparison of exhaust and non-exhaust emissions. Atmos Environ. Feb 1 2019;198:55–69. doi:10.1016/j.atmosenv.2018.10.038

4. Park M, Joo HS, Lee K, et al. Differential toxicities of fine particulate matters from various sources. Sci Rep. Nov 19 2018;8(1):17007. doi:10.1038/s41598-018-35398-0

5. Fu PF, Guo XB, Cheung FMH, Yung KKL. The association between PM2.5 exposure and neurological disorders: A systematic review and meta-analysis. Sci Total Environ. Mar 10 2019;655:1240–1248. doi:10.1016/j.scitotenv.2018.11.218

6. Shou YK, Huang YL, Zhu XZ, Liu CQ, Hu Y, Wang HH. A review of the possible associations between ambient PM2.5 exposures and the development of Alzheimer’s disease. Ecotox Environ Safe. Jun 15 2019;174:344–352. doi:10.1016/j.ecoenv.2019.02.086

7. Alzheimer’s Association. 2022 Alzheimer’s disease facts and figures. Alzheimers Dement. Apr 2022;18(4):700-789. doi:10.1002/alz.12638

8. Brookmeyer R, Abdalla N, Kawas CH, Corrada MM. Forecasting the prevalence of preclinical and clinical Alzheimer’s disease in the United States. Alzheimers Dement. Feb 2018;14(2):121–129. doi:10.1016/j.jalz.2017.10.009

9. Deb A, Thornton JD, Sambamoorthi U, Innes K. Direct and indirect cost of managing alzheimer’s disease and related dementias in the United States. Expert Rev Pharmacoecon Outcomes Res. Apr 2017;17(2):189–202. doi:10.1080/14737167.2017.1313118

10. Killin LO, Starr JM, Shiue IJ, Russ TC. Environmental risk factors for dementia: a systematic review. BMC Geriatr. Oct 12 2016;16(1):175. doi:10.1186/s12877-016-0342-y

11. Kang YJ, Tan HY, Lee CY, Cho H. An Air Particulate Pollutant Induces Neuroinflammation and Neurodegeneration in Human Brain Models. Adv Sci (Weinh). Nov 2021;8(21):e2101251. doi:10.1002/advs.202101251

12. Kilian J, Kitazawa M. The emerging risk of exposure to air pollution on cognitive decline and Alzheimer’s disease - Evidence from epidemiological and animal studies. Biomed J. Jun 2018;41(3):141–162. doi:10.1016/j.bj.2018.06.001

13. Yokoyama AS, Rutledge JC, Medici V. DNA methylation alterations in Alzheimer’s disease. Environ Epigenet. May 2017;3(2):dvx008. doi:10.1093/eep/dvx008

14. Iwata A, Nagata K, Hatsuta H, et al. Altered CpG methylation in sporadic Alzheimer’s disease is associated with APP and MAPT dysregulation. Hum Mol Genet. Feb 1 2014;23(3):648–56. doi:10.1093/hmg/ddt451

15. Wang SC, Oelze B, Schumacher A. Age-specific epigenetic drift in late-onset Alzheimer’s disease. PLoS One. Jul 16 2008;3(7):e2698. doi:10.1371/journal.pone.0002698

16. Smith RG, Hannon E, De Jager PL, et al. Elevated DNA methylation across a 48-kb region spanning the HOXA gene cluster is associated with Alzheimer’s disease neuropathology. Alzheimers & Dementia. Dec 2018;14(12):1580–1588. doi:10.1016/j.jalz.2018.01.017

17. Nicolia V, Cavallaro RA, Lopez-Gonzalez I, et al. DNA Methylation Profiles of Selected Pro-Inflammatory Cytokines in Alzheimer Disease. J Neuropathol Exp Neurol. Jan 1 2017;76(1):27–31. doi:10.1093/jnen/nlw099

18. Wu Y, Qie R, Cheng M, et al. Air pollution and DNA methylation in adults: A systematic review and meta-analysis of observational studies. Environ Pollut. Sep 1 2021;284:117152. doi:10.1016/j.envpol.2021.117152

19. Newcombe EA, Camats-Perna J, Silva ML, Valmas N, Huat TJ, Medeiros R. Inflammation: the link between comorbidities, genetics, and Alzheimer’s disease. J Neuroinflamm. Sep 24 2018;15doi:ARTN 27610.1186/s12974-018-1313-3

20. Fang J, Gao Y, Zhang M, et al. Personal PM(2.5) Elemental Components, Decline of Lung Function, and the Role of DNA Methylation on Inflammation-Related Genes in Older Adults: Results and Implications of the BAPE Study. Environ Sci Technol. Nov 15 2022;56(22):15990–16000. doi:10.1021/acs.est.2c04972

21. Tachibana K, Takayanagi K, Akimoto A, et al. Prenatal diesel exhaust exposure disrupts the DNA methylation profile in the brain of mouse offspring. J Toxicol Sci. Feb 2015;40(1):1–11. doi:10.2131/jts.40.1

22. Wei H, Liang F, Meng G, et al. Redox/methylation mediated abnormal DNA methylation as regulators of ambient fine particulate matter-induced neurodevelopment related impairment in human neuronal cells. Sci Rep. Sep 14 2016;6:33402. doi:10.1038/srep33402

23. Christensen GM, Li Z, Liang D, et al. Fine particulate air pollution and neuropathology markers of Alzheimer’s disease in donors with and without APOE ε4 alleles – results from an autopsy cohort. medRxiv. 2023:2023.04.07.23288288. doi:10.1101/2023.04.07.23288288

24. Besser LM, Kukull WA, Teylan MA, et al. The Revised National Alzheimer’s Coordinating Center’s Neuropathology Form-Available Data and New Analyses. J Neuropathol Exp Neurol. Aug 1 2018;77(8):717–726. doi:10.1093/jnen/nly049

25. Mirra SS, Heyman A, McKeel D, et al. The Consortium to Establish a Registry for Alzheimer’s Disease (CERAD). Part II. Standardization of the neuropathologic assessment of Alzheimer’s disease. Neurology. Apr 1991;41(4):479–86. doi:10.1212/wnl.41.4.479

26. Hyman BT, Phelps CH, Beach TG, et al. National Institute on Aging-Alzheimer’s Association guidelines for the neuropathologic assessment of Alzheimer’s disease. Alzheimers Dement. Jan 2012;8(1):1–13. doi:10.1016/j.jalz.2011.10.007

27. Thal DR, Rub U, Orantes M, Braak H. Phases of A beta-deposition in the human brain and its relevance for the development of AD. Neurology. Jun 25 2002;58(12):1791–800. doi:10.1212/wnl.58.12.1791

28. Zhai X, Russell AG, Sampath P, et al. Calibrating R-LINE model results with observational data to develop annual mobile source air pollutant fields at fine spatial resolution: Application in Atlanta. Atmos Environ. 2016;147:446–457.

29. Sarnat JA, Russell A, Liang D, et al. Developing Multipollutant Exposure Indicators of Traffic Pollution: The Dorm Room Inhalation to Vehicle Emissions (DRIVE) Study. Res Rep Health Eff Inst. Apr 2018;(196):3–75.

30. D’Onofrio D, Kim B, Kim Y, Kim K. Atlanta Roadside Emissions Exposure Study-Methodology & Project Overview. Atlanta Regional Commission; 2016.

31. van Donkelaar A, Hammer MS, Bindle L, et al. Monthly Global Estimates of Fine Particulate Matter and Their Uncertainty. Environ Sci Technol. Nov 16 2021;55(22):15287–15300. doi:10.1021/acs.est.1c05309

32. Stekhoven DJ, Buhlmann P. MissForest--non-parametric missing value imputation for mixed-type data. Bioinformatics. Jan 1 2012;28(1):112–8. doi:10.1093/bioinformatics/btr597

33. Konwar C, Price EM, Wang LQ, Wilson SL, Terry J, Robinson WP. DNA methylation profiling of acute chorioamnionitis-associated placentas and fetal membranes: insights into epigenetic variation in spontaneous preterm births. Epigenetics Chromatin. Oct 29 2018;11(1):63. doi:10.1186/s13072-018-0234-9

34. Johnson WE, Li C, Rabinovic A. Adjusting batch effects in microarray expression data using empirical Bayes methods. Biostatistics. Jan 2007;8(1):118–27. doi:10.1093/biostatistics/kxj037

35. Guintivano J, Aryee MJ, Kaminsky ZA. A cell epigenotype specific model for the correction of brain cellular heterogeneity bias and its application to age, brain region and major depression. Epigenetics. Mar 2013;8(3):290–302. doi:10.4161/epi.23924

36. Aryee MJ, Jaffe AE, Corrada-Bravo H, et al. Minfi: a flexible and comprehensive Bioconductor package for the analysis of Infinium DNA methylation microarrays. Bioinformatics. May 15 2014;30(10):1363–9. doi:10.1093/bioinformatics/btu049

37. Kind AJH, Buckingham WR. Making Neighborhood-Disadvantage Metrics Accessible - The Neighborhood Atlas. N Engl J Med. Jun 28 2018;378(26):2456–2458. doi:10.1056/NEJMp1802313

38. Venables WN, Ripley BD. Modern applied statistics with S-PLUS. Springer Science & Business Media; 2013.

39. Leek JT, Johnson WE, Parker HS, et al. sva: Surrogate variable analysis. R package version. 2019;3(0):882–883.

40. van Iterson M, van Zwet EW, Consortium B, Heijmans BT. Controlling bias and inflation in epigenome-and transcriptome-wide association studies using the empirical null distribution. Genome Biol. Jan 27 2017;18(1):19. doi:10.1186/s13059-016-1131-9

41. Armstrong RA. When to use the Bonferroni correction. Ophthal Physl Opt. Sep 2014;34(5):502–508. doi:10.1111/opo.12131

42. Chadeau-Hyam M, Athersuch TJ, Keun HC, et al. Meeting-in-the-middle using metabolic profiling - a strategy for the identification of intermediate biomarkers in cohort studies. Biomarkers. Feb 2011;16(1):83–8. doi:10.3109/1354750X.2010.533285

43. Zhang H, Zheng Y, Zhang Z, et al. Estimating and testing high-dimensional mediation effects in epigenetic studies. Bioinformatics. Oct 15 2016;32(20):3150–3154. doi:10.1093/bioinformatics/btw351

44. Liu ZH, Shen JC, Barfield R, Schwartz J, Baccarelli AA, Lin XH. Large-Scale Hypothesis Testing for Causal Mediation Effects with Applications in Genome-wide Epigenetic Studies. J Am Stat Assoc. Jan 2 2022;117(537):67–81. doi:10.1080/01621459.2021.1914634

45. Tingley D, Yamamoto T, Hirose K, Keele L, Imai K. mediation: R Package for Causal Mediation Analysis. J Stat Softw. Aug 2014;59(5)

46. Kilanowski A, Merid SK, Abrishamcar S, et al. DNA methylation and aeroallergen sensitization: The chicken or the egg? Clin Epigenetics. Sep 16 2022;14(1):114. doi:10.1186/s13148-022-01332-5

47. Abrishamcar S, Chen JY, Feil D, et al. DNA methylation as a potential mediator of the association between prenatal tobacco and alcohol exposure and child neurodevelopment in a South African birth cohort. Transl Psychiat. Sep 30 2022;12(1)doi:ARTN 418 10.1038/s41398-022-02195-3

48. Imai K, Keele L, Yamamoto T. Identification, inference and sensitivity analysis for causal mediation effects. 2010;

49. Imai K, Keele L, Tingley D, Yamamoto T. Causal mediation analysis using R. Springer; 2010:129–154.

50. Maksimovic J, Oshlack A, Phipson B. Gene set enrichment analysis for genome-wide DNA methylation data. Genome Biology. Jun 8 2021;22(1)doi:ARTN 173 10.1186/s13059-021-02388-x

51. Hansen K. IlluminaHumanMethylationEPICanno. ilm10b2. hg19: annotation for Illumina’s EPIC methylation arrays. R package version 0.6. 0. 2016.

52. Battram T, Yousefi P, Crawford G, et al. The EWAS Catalog: a database of epigenome-wide association studies. Wellcome Open Res. 2022;7:41. doi:10.12688/wellcomeopenres.17598.2

53. Eisenberg DT, Kuzawa CW, Hayes MG. Worldwide allele frequencies of the human apolipoprotein E gene: climate, local adaptations, and evolutionary history. Am J Phys Anthropol. Sep 2010;143(1):100–11. doi:10.1002/ajpa.21298

54. Sordillo JE, Cardenas A, Qi C, et al. Residential PM(2.5) exposure and the nasal methylome in children. Environ Int. Aug 2021;153:106505. doi:10.1016/j.envint.2021.106505

55. Yu S, Dai J, Ma M, et al. RBCK1 promotes p53 degradation via ubiquitination in renal cell carcinoma. Cell Death Dis. Mar 15 2019;10(4):254. doi:10.1038/s41419-019-1488-2

56. Jazvinscak Jembrek M, Slade N, Hof PR, Simic G. The interactions of p53 with tau and Ass as potential therapeutic targets for Alzheimer’s disease. Prog Neurobiol. Sep 2018;168:104–127. doi:10.1016/j.pneurobio.2018.05.001

57. Taminiau A, Draime A, Tys J, et al. HOXA1 binds RBCK1/HOIL-1 and TRAF2 and modulates the TNF/NF-kappaB pathway in a transcription-independent manner. Nucleic Acids Res. Sep 6 2016;44(15):7331–49. doi:10.1093/nar/gkw606

58. Srinivasan M, Lahiri DK. Significance of NF-kappaB as a pivotal therapeutic target in the neurodegenerative pathologies of Alzheimer’s disease and multiple sclerosis. Expert Opin Ther Targets. Apr 2015;19(4):471–87. doi:10.1517/14728222.2014.989834

59. Snow WM, Albensi BC. Neuronal Gene Targets of NF-kappaB and Their Dysregulation in Alzheimer’s Disease. Front Mol Neurosci. 2016;9:118. doi:10.3389/fnmol.2016.00118

60. Caccamo A, Branca C, Piras IS, et al. Necroptosis activation in Alzheimer’s disease. Nat Neurosci. Sep 2017;20(9):1236–1246. doi:10.1038/nn.4608

61. Jayaraman A, Htike TT, James R, Picon C, Reynolds R. TNF-mediated neuroinflammation is linked to neuronal necroptosis in Alzheimer’s disease hippocampus. Acta Neuropathol Commun. Sep 28 2021;9(1):159. doi:10.1186/s40478-021-01264-w

62. Wang B, Bao S, Zhang Z, et al. A rare variant in MLKL confers susceptibility to ApoE varepsilon4-negative Alzheimer’s disease in Hong Kong Chinese population. Neurobiol Aging. Aug 2018;68:160 e1–160 e7. doi:10.1016/j.neurobiolaging.2018.03.006

63. Chen S, Liu H, Wang S, et al. The Neuroprotection of Verbascoside in Alzheimer’s Disease Mediated through Mitigation of Neuroinflammation via Blocking NF-kappaB-p65 Signaling. Nutrients. Mar 29 2022;14(7)doi:10.3390/nu14071417

64. Lee JH, Cheng R, Vardarajan B, et al. Genetic Modifiers of Age at Onset in Carriers of the G206A Mutation in PSEN1 With Familial Alzheimer Disease Among Caribbean Hispanics. JAMA Neurol. Sep 2015;72(9):1043–51. doi:10.1001/jamaneurol.2015.1424

65. Chao MW, Yang CH, Lin PT, et al. Exposure to PM(2.5) causes genetic changes in fetal rat cerebral cortex and hippocampus. Environ Toxicol. Apr 2017;32(4):1412–1425. doi:10.1002/tox.22335

66. Vdovenko D, Bachmann M, Wijnen WJ, Hottiger MO, Eriksson U, Valaperti A. The adaptor protein c-Cbl-associated protein (CAP) limits pro-inflammatory cytokine expression by inhibiting the NF-kappaB pathway. Int Immunopharmacol. Oct 2020;87:106822. doi:10.1016/j.intimp.2020.106822

67. Heckman PR, Wouters C, Prickaerts J. Phosphodiesterase inhibitors as a target for cognition enhancement in aging and Alzheimer’s disease: a translational overview. Curr Pharm Des. 2015;21(3):317–31. doi:10.2174/1381612820666140826114601

68. Puthiyedth N, Riveros C, Berretta R, Moscato P. Identification of Differentially Expressed Genes through Integrated Study of Alzheimer’s Disease Affected Brain Regions. PLoS One. 2016;11(4):e0152342. doi:10.1371/journal.pone.0152342

69. Iwakiri M, Mizukami K, Ikonomovic MD, et al. Changes in hippocampal GABABR1 subunit expression in Alzheimer’s patients: association with Braak staging. Acta Neuropathol. May 2005;109<otherinfo>(5)</otherinfo>:467-74. doi:10.1007/s00401-005-0985-9

70. Shang Y, Das S, Rabold R, Sham JS, Mitzner W, Tang WY. Epigenetic alterations by DNA methylation in house dust mite-induced airway hyperresponsiveness. Am J Respir Cell Mol Biol. Aug 2013;49(2):279–87. doi:10.1165/rcmb.2012-0403OC

71. Oddo S. The role of mTOR signaling in Alzheimer disease. Front Biosci (Schol Ed). Jan 1 2012;4(3):941–52. doi:10.2741/s310

72. Vineis P, van Veldhoven K, Chadeau-Hyam M, Athersuch TJ. Advancing the application of omics-based biomarkers in environmental epidemiology. Environ Mol Mutagen. Aug 2013;54(7):461–7. doi:10.1002/em.21764

73. Wilson JG, Kingham S, Pearce J, Sturman AP. A review of intraurban variations in particulate air pollution: Implications for epidemiological research. Atmos Environ. Nov 2005;39(34):6444–6462. doi:10.1016/j.atmosenv.2005.07.030

74. Dominici F, Peng RD, Barr CD, Bell ML. Protecting human health from air pollution: shifting from a single-pollutant to a multipollutant approach. Epidemiology. Mar 2010;21(2):187–94. doi:10.1097/EDE.0b013e3181cc86e8

75. Alemany S, Crous-Bou M, Vilor-Tejedor N, et al. Associations between air pollution and biomarkers of Alzheimer’s disease in cognitively unimpaired individuals. Environment International. Dec 2021;157doi:ARTN 106 86410.1016/j.envint.2021.106864

