## Supplementary material for "Differential DNA Methylation in the Brain as Potential Mediator of the Association between Traffic-related PM_2.5_ and Neuropathology Markers of Alzheimer’s Disease": ADRC_DNAm_suppl_submit.docx

**Methods**

*Genome-wide DNA methylation quality control and processing*

All DNAm data were preprocessed to identify low-quality samples, exclude specific probes, and reduce the impact of batch effects. Raw intensity files were converted to methylation beta values ranging on a continuous scale from 0 to 1 for each of the CpG sites measured on the array. The Illumina’s 636 control probes were used via the R package *ewastools* to assess technique parameters including array staining, extension, hybridization, target removal, specificity, and bisulfite conversion.^1^ Additional sample outlier detection was implemented based on detection *p* value, beadcount, and distance from the group average in principal components. All 167 samples passed the aforementioned checks. Then, participant sex and replicate status were also verified using XY probes and SNP probes, respectively. One sample with discordant sex compared to the survey data was excluded, and 6 replicates were also excluded. One sample was removed due to potential contamination by another sample. As a result, one hundred and fifty-nine samples remained. Then, the Funnorm function and Combat function was used to normalize the distributions to reduce technical variation and correct for differences between type I and type II probe signals. The following probes were further removed: XY probes, low-quality probes with missing in more than 5% of samples, probes with poor detection p-values, probes predicted to cross-hybridize, probes that bind to the sex chromosomes, polymorphic probes, and probes with infinite values. In total, after all preprocessing steps, 159 samples and 789,286 CpG sites remained for the down-stream analysis. We used estimateCellCounts function in the R package *minfi* to obtain the cell-type proportions (neuronal vs. non-neuronal cells) using the reference dataset published previously.^2,3^

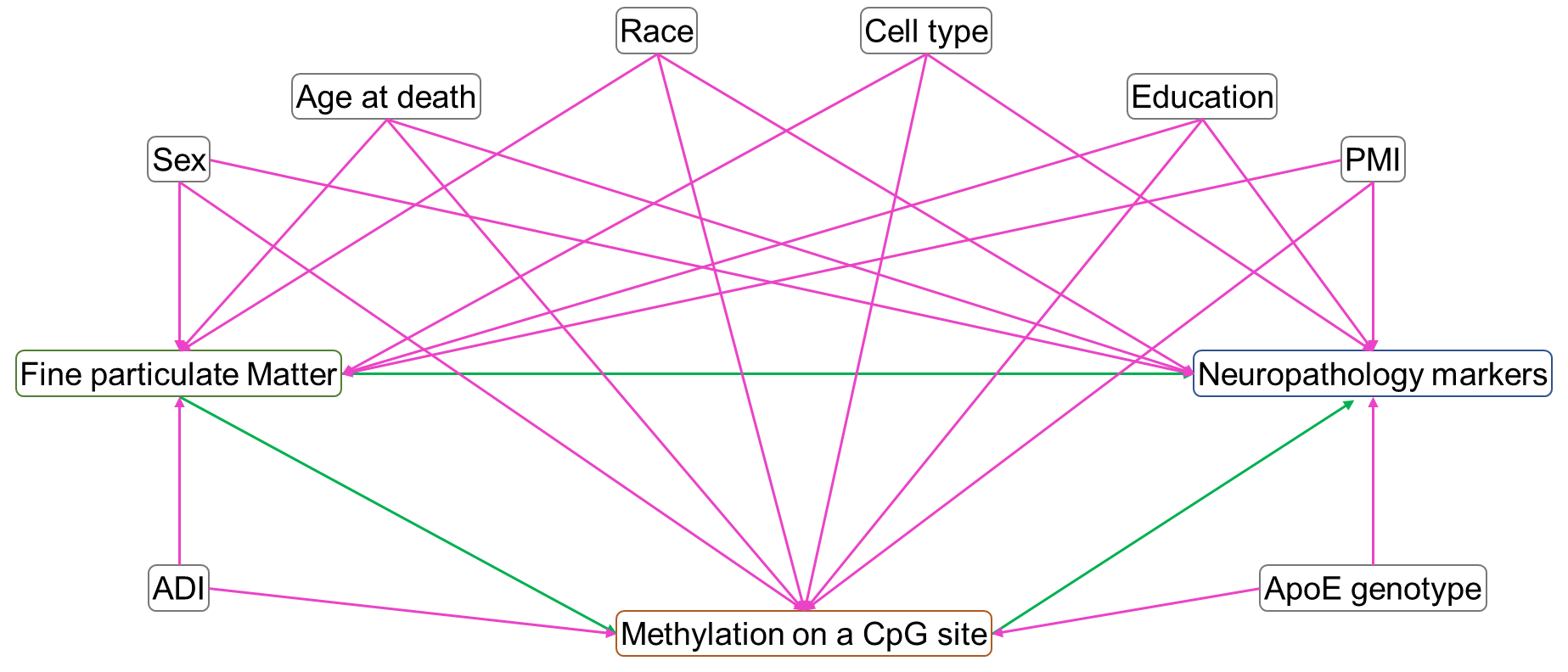

**Figure S1.** Directed acyclic graph of the confounding structure for the association between fine particulate matter exposure and neuropathology markers. PMI, post-mortem interval; ADI, area deprivation index; ApoE, Apolipoprotein E.

**Table S1.** The overlapping CpGs associated with both traffic-related PM_2.5_ exposure prior to death and neuropathology markers.

| **CpG** | **chr** | **Position** | **Gene** |  | **Coefficients ^a^** | ***p*-values** |
| --- | --- | --- | --- | --- | --- | --- |
| **A. CpGs with PM_2.5_ exposures** | | |  |  |  |  |
| cg16342341 | 4 | 186,571,988 | *SORBS2* | 1-year exposure | 0.0102 | 1.27×10^-3^ |
| cg01835635 | 11 | 116,693,535 | *APOA4* | 1-year exposure | -0.0168 | 6.16×10^-4^ |
|  |  |  |  | 3-year exposure | -0.0155 | 1.29×10^-3^ |
| cg09830308 | 16 | 74,734,321 | *MLKL* | 1-year exposure | 0.0163 | 1.15×10^-3^ |
|  |  |  |  | 3-year exposure | 0.0161 | 9.44×10^-4^ |
|  |  |  |  | 5-year exposure | 0.0151 | 8.47×10^-4^ |
| cg27459981 | 16 | 74,734,571 | *MLKL* | 3-year exposure | 0.0155 | 1.11×10^-3^ |
|  |  |  |  | 5-year exposure | 0.0142 | 1.37×10^-3^ |
| **B. CpGs with neuropathology markers** | | |  |  |  |  |
| cg16342341 | 4 | 186,571,988 | *SORBS2* | CERAD | 0.52 | 6.03×10^-4^ |
|  |  |  |  | ABC | 0.44 | 5.37×10^-4^ |
| cg01835635 | 11 | 116,693,535 | *APOA4* | CERAD | -0.63 | 4.36×10^-4^ |
| cg09830308 | 16 | 74,734,321 | *MLKL* | Braak stage | -0.51 | 3.09×10^-4^ |
| cg27459981 | 16 | 74,734,571 | *MLKL* | Braak stage | -0.82 | 1.70×10^-5^ |

Abbreviations: PM_2.5_, fine particulate matter; chr, chromosome; *SORBS2*, sorbin and SH3 domain-containing protein 2; *APOA4*, Apolipoprotein A-IV; *MLKL*, mixed lineage kinase domain like pseudokinase.

^a^ The coefficients for PM_2.5_ exposures represent the change in the beta values of CpG sites associated with one-unit increase in the exposures; the coefficients for neuropathology markers represent the change in the neuropathology markers associated with one-interquartile-range increase in the beta values of CpG sites.

**Table S2.** Indirect effect estimated by causal mediation analysis via the R package *mediation* of CpG sites selected by high-dimensional mediation analysis for the associations of PM_2.5_ exposure with Braak stage and ABC score ^a^.

| **CpG** | **chr** | **Gene** | **Exposure** | **Outcome** | **ACME ^c^** | **Total effect ^d^** |
| --- | --- | --- | --- | --- | --- | --- |
| cg09579061 | 1 | *ADORA1* | 1-year | Braak | -0.183 (-0.348, -5.00E-02) | 0.032 (-0.318, 4.30E-01) |
| cg23932332 | 1 | *DUSP10* | 3-year | ABC | 0.056 (0.005, 1.50E-01) | 0.086 (-0.110, 2.80E-01) |
|  |  |  | 5-year | ABC | 0.060 (0.002, 1.70E-01) | 0.104 (-0.081, 3.10E-01) |
| cg08512806 | 1 | *TARBP1* | 3-year | ABC | 0.058 (0.008, 1.30E-01) | 0.084 (-0.107, 3.00E-01) |
|  |  |  | 5-year | ABC | 0.063 (0.009, 1.30E-01) | 0.102 (-0.080, 3.10E-01) |
| cg10705045 | 2 | *RNF144A* | 5-year | ABC | 0.063 (0.001, 1.40E-01) | 0.109 (-0.079, 3.10E-01) |
| cg17275287 | 2 | Intergenic | 3-year | ABC | 0.085 (0.019, 1.70E-01) | 0.079 (-0.118, 3.00E-01) |
|  |  |  | 5-year | ABC | 0.089 (0.020, 1.80E-01) | 0.097 (-0.093, 3.00E-01) |
| cg05532414 | 2 | Intergenic | 3-year | ABC | 0.071 (0.004, 1.70E-01) | 0.090 (-0.084, 3.20E-01) |
| cg07963191 | 2 | *PDE11A* | 3-year | ABC | 0.061 (0.005, 1.40E-01) | 0.080 (-0.103, 3.00E-01) |
| cg26109897 | 4 | *TBC1D14* | 3-year | ABC | 0.085 (0.010, 1.90E-01) | 0.090 (-0.098, 3.10E-01) |
| cg26877022 | 4 | *POLR2B* | 3-year | ABC | 0.080 (0.015, 1.80E-01) | 0.089 (-0.092, 3.10E-01) |
|  |  |  | 5-year | ABC | 0.077 (0.011, 1.80E-01) | 0.107 (-0.079, 3.10E-01) |
| cg16342341 | 4 | *SORBS2* | 1-year | ABC | 0.097 (0.021, 1.80E-01) | 0.034 (-0.168, 2.30E-01) |
|  |  |  | 3-year | ABC | 0.076 (0.017, 1.60E-01) | 0.080 (-0.106, 2.80E-01) |
|  |  |  | 5-year | ABC | 0.078 (0.017, 1.50E-01) | 0.098 (-0.093, 3.20E-01) |
| cg17444747 | 5 | *COL23A1* | 5-year | ABC | 0.074 (0.015, 1.50E-01) | 0.098 (-0.085, 2.90E-01) |
| cg27297993 | 6 | *GABBR1* | 3-year | ABC | 0.064 (0.009, 1.40E-01) | 0.084 (-0.091, 3.00E-01) |
|  |  |  | 5-year | ABC | 0.066 (0.003, 1.40E-01) | 0.103 (-0.076, 3.00E-01) |
| cg00829961 | 8 | Intergenic | 3-year | ABC | 0.075 (0.009, 1.70E-01) | 0.092 (-0.092, 3.10E-01) |
|  |  |  | 5-year | ABC | 0.075 (0.012, 1.70E-01) | 0.110 (-0.078, 3.30E-01) |
| cg02987635 | 10 | *C10orf11* | 3-year | ABC | 0.063 (0.004, 1.50E-01) | 0.079 (-0.099, 3.00E-01) |
| cg06805557 | 11 | *APBB1* | 5-year | ABC | 0.062 (0.007, 1.30E-01) | 0.101 (-0.104, 3.00E-01) |
| cg19969778 | 11 | *SIAE;*  *SPA17* | 3-year | ABC | 0.065 (0.008, 1.30E-01) | 0.080 (-0.108, 3.10E-01) |
|  |  |  | 5-year | ABC | 0.063 (0.010, 1.30E-01) | 0.098 (-0.092, 3.10E-01) |
| cg17562250 | 14 | *LOC101927124;*  *HEATR5A* | 1-year | ABC | -0.077 (-0.174, -1.00E-02) | 0.039 (-0.169, 2.60E-01) |
|  |  |  | 3-year | ABC | -0.065 (-0.169, 0.00E+00) | 0.085 (-0.099, 3.00E-01) |
| cg20713102 | 15 | *ZSCAN2* | 5-year | ABC | 0.074 (0.014, 1.60E-01) | 0.106 (-0.083, 3.10E-01) |
| cg09088153 | 15 | Intergenic | 3-year | ABC | 0.072 (0.013, 1.50E-01) | 0.089 (-0.094, 3.20E-01) |
| cg27181554 | 16 | *SEPX1* | 1-year | ABC | 0.084 (0.021, 1.80E-01) | 0.039 (-0.162, 2.70E-01) |
|  |  |  | 3-year | ABC | 0.069 (0.015, 1.50E-01) | 0.085 (-0.108, 2.80E-01) |
| cg20389589 | 16 | *FAM57B* | 3-year | ABC | 0.069 (0.003, 1.60E-01) | 0.084 (-0.120, 2.90E-01) |
| cg06832209 | 16 | *ADGRG3* | 3-year | ABC | 0.078 (0.015, 1.60E-01) | 0.089 (-0.101, 2.80E-01) |
| cg09830308 | 16 | *MLKL* | 1-year | ABC | -0.072 (-0.161, -1.00E-02) | 0.030 (-0.171, 2.40E-01) |
|  |  |  | 1-year | Braak | -0.152 (-0.319, -1.00E-02) | 0.023 (-0.318, 3.80E-01) |
|  |  |  | 3-year | ABC | -0.071 (-0.151, 0.00E+00) | 0.073 (-0.130, 2.90E-01) |
|  |  |  | 5-year | ABC | -0.073 (-0.163, -1.00E-02) | 0.092 (-0.115, 3.10E-01) |
| cg27459981 | 16 | *MLKL* | 1-year | ABC | -0.089 (-0.178, -2.00E-02) | 0.028 (-0.198, 2.50E-01) |
|  |  |  | 1-year | Braak | -0.182 (-0.364, -4.00E-02) | 0.019 (-0.333, 3.60E-01) |
|  |  |  | 3-year | ABC | -0.086 (-0.168, -2.00E-02) | 0.071 (-0.132, 2.80E-01) |
|  |  |  | 5-year | ABC | -0.087 (-0.179, -2.00E-02) | 0.089 (-0.093, 2.90E-01) |
| cg08591058 | 17 | Intergenic | 1-year | ABC | -0.092 (-0.175, -2.00E-02) | 0.030 (-0.162, 2.50E-01) |
|  |  |  | 3-year | ABC | -0.084 (-0.165, -2.00E-02) | 0.073 (-0.125, 2.90E-01) |
| cg26535871 | 17 | *MGAT5B* | 3-year | ABC | -0.063 (-0.147, 0.00E+00) | 0.070 (-0.129, 2.80E-01) |
|  |  |  | 5-year | ABC | -0.065 (-0.164, -1.00E-02) | 0.089 (-0.114, 2.90E-01) |
| cg00633834 | 19 | *AKT1S1;*  *TBC1D17* | 5-year | ABC | 0.081 (0.017, 1.60E-01) | 0.095 (-0.090, 2.90E-01) |

Abbreviations: PM_2.5_, fine particulate matter; chr, chromosome; ACME, average causal mediated effect (i.e., indirect effect).

^a^ All CpG sites that were selected by *DACT* and had a significant ACME were associated with Braak stage or ABC score.

^b^ The *p*-values of mediation effect testing conducted by *DACT*.

^c^ The ACME was associated with one-interquartile-range increase in beta values of CpG sites.

^d^ Effect estimates, associated with 1-unit increase, of PM_2.5_ exposures on neuropathology markers.

**Table S3.** The top 10 KEGG pathways enriched by differentially methylated CpG sites that were associated with PM2.5 exposure and neuropathology markers, respectively.

| **Exposure/**  **outcome** | **KEGG terms** | **Description** | **N of genes in the KEGG term** | **N of differentially methylated genes** | ***p*-value** | **False discovery rate** | **Significant genes** |
| --- | --- | --- | --- | --- | --- | --- | --- |
| 1-year exposure | path:hsa04137 | Mitophagy - animal | 69 | 8 | 0.0106011 | 1 | CSNK2B, MRAS, KRAS, NRAS, PRKN, AMBRA1, MAPK9, ATG5, USP15 |
|  | path:hsa00030 | Pentose phosphate pathway | 25 | 4 | 0.0145971 | 1 | GPI, PGD, TALDO1, H6PD |
|  | path:hsa04935 | Growth hormone synthesis, secretion and action | 117 | 13 | 0.0168878 | 1 | GHR, GNAI2, IRS1, KRAS, MAP3K1, NRAS, PRKCB, MAPK9, MAP2K1, SSTR3, CACNA1C, IRS2, BCAR1 |
|  | path:hsa04140 | Autophagy - animal | 134 | 13 | 0.0175384 | 1 | EIF2S1, ERN1, MRAS, IRS1, KRAS, NRAS, SH3GLB1, ATG16L1, AMBRA1, MAPK9, MAP2K1, RPTOR, IRS2, ATG5 |
|  | path:hsa05010 | Alzheimer disease | 359 | 23 | 0.0278593 | 1 | APC2, COX7C, CSNK2B, DVL3, EIF2S1, ERN1, ATF6, GRIN2B, APP, IRS1, KRAS, LRP1, LRP5, NRAS, ATP5PO, AMBRA1, MAPK9, MAP2K1, PSMD13, WNT6, CACNA1C, AXIN1, IRS2, SLC39A13, COX7A2L |
|  | path:hsa04720 | Long-term potentiation | 64 | 8 | 0.0319516 | 1 | GRIA1, GRIN2B, KRAS, NRAS, PRKCB, MAP2K1, RAP1B, CACNA1C |
|  | path:hsa04144 | Endocytosis | 244 | 19 | 0.0365904 | 1 | EHD1, AGAP2, AP2A2, PSD3, PIP5K1C, LDLRAP1, CHMP4A, HSPA1A, HSPA1L, SH3GLB1, RUFY2, VPS35, PARD3, RAB5A, SH3GL1, SH3GL2, EEA1, PARD6G, RAB11A, ASAP2, IQSEC1 |
|  | path:hsa01200 | Carbon metabolism | 105 | 8 | 0.0463284 | 1 | SDSL, PHGDH, GPI, PCCA, PGD, TALDO1, SUCLG2, H6PD |
|  | path:hsa04213 | Longevity regulating pathway - multiple species | 61 | 7 | 0.0484809 | 1 | HSPA1A, HSPA1L, IRS1, KRAS, NRAS, RPTOR, IRS2, ATG5 |
|  | path:hsa05034 | Alcoholism | 165 | 11 | 0.0489502 | 1 | H4C16, GNAI2, GRIN2B, KRAS, NRAS, MAP2K1, H2AC14, H2AC16, H2BC14, H3C10, H4C3, HDAC4 |
| 3-year exposure | path:hsa04930 | Type II diabetes mellitus | 45 | 8 | 0.0045222 | 0.7312499 | HK1, IRS1, PRKCZ, MAPK9, SLC2A4, CACNA1B, CACNA1C, IRS2 |
|  | path:hsa04213 | Longevity regulating pathway - multiple species | 61 | 9 | 0.0052512 | 0.7312499 | ADCY1, HSPA1A, HSPA1L, IRS1, KRAS, NRAS, PRKAG2, RPTOR, IRS2, ATG5 |
|  | path:hsa00900 | Terpenoid backbone biosynthesis | 23 | 4 | 0.0062322 | 0.7312499 | ZMPSTE24, FNTA, ICMT, IDI1 |
|  | path:hsa04720 | Long-term potentiation | 64 | 9 | 0.0103579 | 0.9114961 | ADCY1, GRIA1, GRIN2B, KRAS, NRAS, PRKCB, MAP2K1, RAP1B, CACNA1C |
|  | path:hsa04650 | Natural killer cell mediated cytotoxicity | 119 | 10 | 0.0180657 | 1 | HCST, PTK2B, FYN, KRAS, NFATC2, NRAS, PAK1, PRKCB, MAP2K1, VAV2 |
|  | path:hsa04211 | Longevity regulating pathway | 88 | 10 | 0.0235191 | 1 | ADCY1, EHMT2, CREB1, SESN3, IRS1, KRAS, NRAS, PRKAG2, RPTOR, IRS2, ATG5 |
|  | path:hsa05010 | Alzheimer disease | 359 | 23 | 0.0287495 | 1 | APC2, COX7C, DVL3, EIF2S1, ATF6, GRIN2B, APP, IRS1, KRAS, LRP1, LRP5, ATP2A2, NRAS, ATP5PO, AMBRA1, MAPK9, MAP2K1, PSMA2, PSMD13, SLC39A8, WNT6, CACNA1C, AXIN1, CASP8, IRS2 |
|  | path:hsa04910 | Insulin signaling pathway | 131 | 12 | 0.0312015 | 1 | HK1, ACACA, IRS1, KRAS, NRAS, PRKAG2, PRKCZ, MAPK9, MAP2K1, RPTOR, SLC2A4, IRS2 |
|  | path:hsa04144 | Endocytosis | 244 | 19 | 0.032739 | 1 | EHD1, AP2A2, PSD3, PIP5K1C, CYTH4, CHMP4A, HSPA1A, HSPA1L, SH3GLB1, VTA1, RUFY2, PRKCZ, PARD3, RAB5A, WIPF3, SH3GL1, EEA1, RAB11A, DNAJC6, IQSEC1 |
|  | path:hsa04935 | Growth hormone synthesis, secretion and action | 117 | 12 | 0.0330168 | 1 | ADCY1, CREB1, IRS1, KRAS, MAP3K1, NRAS, PRKCB, MAPK9, MAP2K1, SSTR3, CACNA1C, IRS2 |
| 5-year exposure | path:hsa00590 | Arachidonic acid metabolism | 61 | 7 | 0.0030161 | 1 | PLB1, ALOX5, GGT1, PLA2G4D, GPX4, ALOXE3, PLA2G6 |
|  | path:hsa04720 | Long-term potentiation | 64 | 9 | 0.0134991 | 1 | ADCY1, GRIA1, GRIN2B, NRAS, PPP3CA, PRKCB, MAP2K1, RAP1B, CACNA1C |
|  | path:hsa04930 | Type II diabetes mellitus | 45 | 7 | 0.0195656 | 1 | HK1, PRKCZ, MAPK9, SLC2A4, CACNA1B, CACNA1C, IRS2 |
|  | path:hsa04931 | Insulin resistance | 105 | 11 | 0.021487 | 1 | PPARGC1B, CREB1, SLC27A1, NOS3, PRKAG2, PRKCB, PRKCZ, MAPK9, SLC2A4, IRS2, CREB5 |
|  | path:hsa00450 | Selenocompound metabolism | 17 | 3 | 0.0343345 | 1 | TXNRD2, PSTK, PAPSS1 |
|  | path:hsa00513 | Various types of N-glycan biosynthesis | 38 | 5 | 0.0400335 | 1 | HEXB, STT3A, ALG11, MAN1C1, CHST8, TUSC3 |
|  | path:hsa04370 | VEGF signaling pathway | 59 | 7 | 0.0412512 | 1 | PLA2G4D, NFATC2, NOS3, NRAS, PPP3CA, PRKCB, MAP2K1 |
|  | path:hsa00900 | Terpenoid backbone biosynthesis | 23 | 3 | 0.0437082 | 1 | ZMPSTE24, FNTA, ICMT |
|  | path:hsa04136 | Autophagy - other | 30 | 4 | 0.0537717 | 1 | WIPI2, ATG16L1, RPTOR, ATG5 |
|  | path:hsa00591 | Linoleic acid metabolism | 30 | 3 | 0.0537983 | 1 | PLB1, PLA2G4D, PLA2G6 |
| Braak stage | path:hsa05032 | Morphine addiction | 88 | 13 | 0.0011824 | 0.4161928 | ADCY1, PDE10A, ADCY5, ADCY9, ADCY4, GNG7, GNGT2, GRK5, ARRB1, PDE11A, PDE1A, PDE3B, PDE4D, PRKCA |
|  | path:hsa05110 | Vibrio cholerae infection | 49 | 7 | 0.0038668 | 0.6710898 | ADCY9, ARF1, KCNQ1, ATP6V0C, PLCG2, PRKCA, TJP1, ATP6V0D1 |
|  | path:hsa04261 | Adrenergic signaling in cardiomyocytes | 148 | 15 | 0.0074374 | 0.6710898 | CACNG3, ADCY1, ADCY5, ADCY9, CREB1, CREM, ADCY4, KCNQ1, PPP1CA, PPP2R1A, PRKCA, TPM3, CACNA1C, CACNB3, CACNA2D2 |
|  | path:hsa05130 | Pathogenic Escherichia coli infection | 189 | 15 | 0.0081416 | 0.6710898 | WIPF2, TUBB, CYFIP1, CYTH4, IKBKB, ARF1, MYO1C, MYO5C, BAIAP2L1, RAC1, WIPF3, TJP1, TNFRSF1A, MYH14, TNFRSF10A |
|  | path:hsa04015 | Rap1 signaling pathway | 207 | 19 | 0.0095325 | 0.6710898 | ADCY1, ADCY5, ADCY9, CSF1R, CTNND1, ADCY4, EPHA2, FGF6, FLT4, FYB1, PRKD2, RAPGEF1, NGFR, EVL, PRKCA, PARD3, RAC1, VEGFC, FGF23, FGF18 |
|  | path:hsa04061 | Viral protein interaction with cytokine and cytokine receptor | 97 | 6 | 0.0146182 | 0.8213017 | CSF1R, CX3CR1, IL6R, TNFRSF1A, TNFRSF10A, IL18R1 |
|  | path:hsa00230 | Purine metabolism | 123 | 11 | 0.0163327 | 0.8213017 | ADCY1, PDE10A, ADCY5, ADCY9, ADCY4, AK4, AMPD3, GUCY2D, PDE11A, PDE1A, PDE3B, PDE4D |
|  | path:hsa04151 | PI3K-Akt signaling pathway | 339 | 24 | 0.0215544 | 0.849279 | PIK3AP1, COL1A1, CREB1, CSF1R, EPHA2, FGF6, PHLPP2, FLT4, GNG7, GNGT2, IKBKB, IL6R, LAMA3, MDM2, NGFR, NOS3, PPP2R1A, PRKCA, RAC1, CCND1, TNXB, VEGFC, FGF23, FGF18, CCND3 |
|  | path:hsa04010 | MAPK signaling pathway | 283 | 22 | 0.0217145 | 0.849279 | CACNG3, CSF1R, DUSP4, EPHA2, FGF6, FLT4, IKBKB, ARRB1, NGFR, PRKCA, RAC1, RASGRF1, RPS6KA2, TNFRSF1A, VEGFC, CACNA1C, CACNB3, FGF23, MKNK1, FGF18, CACNA2D2, CD14 |
|  | path:hsa04014 | Ras signaling pathway | 227 | 18 | 0.0255507 | 0.8993838 | CSF1R, EPHA2, FGF6, RASA3, RGL1, FLT4, GNG7, GNGT2, IKBKB, NGFR, PLCG2, PRKCA, RGL2, RAC1, RASGRF1, VEGFC, FGF23, FGF18, KSR1 |
| CERAD | path:hsa05130 | Pathogenic Escherichia coli infection | 189 | 18 | 0.0009299 | 0.3273211 | TAB2, CYFIP1, GAPDH, CYFIP2, IL6, ARF1, MYO1A, MYO1C, MYO5A, MYO5B, NFKB1, CLDN18, BAIAP2L1, MAPK10, RAC1, TNFRSF1A, TRAF2, TUBB6 |
|  | path:hsa01523 | Antifolate resistance | 29 | 5 | 0.00559 | 0.8349026 | ABCC4, IL6, NFKB1, SHMT1, TYMS, ABCG2 |
|  | path:hsa05146 | Amoebiasis | 98 | 10 | 0.0131476 | 0.8349026 | LAMB4, LAMA1, IL6, ITGAM, NFKB1, NOS2, PRKCA, PTK2, ACTN4, ACTN1 |
|  | path:hsa05131 | Shigellosis | 238 | 18 | 0.0143556 | 0.8349026 | RBCK1, TAB2, ARF1, MDM2, NFKB1, WIPI1, MAPK10, PTK2, RPTOR, RAC1, ELMO2, TNFRSF1A, TRAF2, ACTN4, TLN2, AKT1S1, ACTN1, ATG5 |
|  | path:hsa00430 | Taurine and hypotaurine metabolism | 16 | 3 | 0.0149302 | 0.8349026 | FMO1, GGT7, BAAT |
|  | path:hsa05171 | Coronavirus disease - COVID-19 | 215 | 13 | 0.0162667 | 0.8349026 | ADAR, RPL22L1, TAB2, RPL36, IFNAR1, IL6, NFKB1, PRKCA, MAPK10, RPL18, RPL26, STAT2, TNFRSF1A, TRAF3 |
|  | path:hsa04622 | RIG-I-like receptor signaling pathway | 65 | 6 | 0.0166032 | 0.8349026 | ADAR, NFKB1, MAPK10, TRAF2, TRAF3, ATG5 |
|  | path:hsa03060 | Protein export | 23 | 3 | 0.0343889 | 1 | SEC61A1, SEC61A2, SEC11C |
|  | path:hsa05132 | Salmonella infection | 240 | 16 | 0.0352334 | 1 | ACTR1B, TAB2, CYFIP1, PLEKHM2, GAPDH, CYFIP2, IL6, ARF1, NFKB1, DCTN4, MAPK10, RAC1, ELMO2, TNFRSF1A, TRAF2, TUBB6 |
|  | path:hsa04211 | Longevity regulating pathway | 88 | 9 | 0.0370466 | 1 | ADCY3, ADCY5, NFKB1, PRKAG2, RPTOR, TSC2, EHMT1, ULK1, AKT1S1, ATG5 |
| ABC | path:hsa04672 | Intestinal immune network for IgA production | 44 | 5 | 0.0102832 | 1 | HLA-DOA, IL6, IL15, CD28, CD40 |
|  | path:hsa04940 | Type I diabetes mellitus | 41 | 5 | 0.0141776 | 1 | HLA-B, HLA-DOA, PTPRN, PTPRN2, CD28 |
|  | path:hsa05330 | Allograft rejection | 34 | 4 | 0.018464 | 1 | HLA-B, HLA-DOA, CD28, CD40 |
|  | path:hsa05332 | Graft-versus-host disease | 38 | 4 | 0.0199458 | 1 | HLA-B, HLA-DOA, IL6, CD28 |
|  | path:hsa00740 | Riboflavin metabolism | 8 | 2 | 0.0207243 | 1 | ENPP1, ACP2 |
|  | path:hsa05320 | Autoimmune thyroid disease | 46 | 4 | 0.0371603 | 1 | HLA-B, HLA-DOA, CD28, CD40 |
|  | path:hsa00130 | Ubiquinone and other terpenoid-quinone biosynthesis | 11 | 2 | 0.0407866 | 1 | HPD, COQ5 |
|  | path:hsa05131 | Shigellosis | 238 | 16 | 0.0428873 | 1 | RBCK1, TAB2, MTOR, ITPR1, ARF1, MDM2, NFKB1, PLCG2, PTK2, RPTOR, RAC1, TNFRSF1A, C3, ACTN4, AKT1S1, ACTN1 |
|  | path:hsa05150 | Staphylococcus aureus infection | 90 | 5 | 0.0528039 | 1 | CFH, HLA-DOA, KRT9, SELPLG, C3 |
|  | path:hsa04330 | Notch signaling pathway | 58 | 6 | 0.0547066 | 1 | DTX3L, DTX1, HEYL, NOTCH2, ADAM17, TLE2, NCOR2 |

**References**

1.	Murat K, Gruning B, Poterlowicz PW, Westgate G, Tobin DJ, Poterlowicz K. Ewastools: Infinium Human Methylation BeadChip pipeline for population epigenetics integrated into Galaxy. *Gigascience*. May 1 2020;9(5)doi:10.1093/gigascience/giaa049

2.	Guintivano J, Aryee MJ, Kaminsky ZA. A cell epigenotype specific model for the correction of brain cellular heterogeneity bias and its application to age, brain region and major depression. *Epigenetics*. Mar 2013;8(3):290-302. doi:10.4161/epi.23924

3.	Aryee MJ, Jaffe AE, Corrada-Bravo H, et al. Minfi: a flexible and comprehensive Bioconductor package for the analysis of Infinium DNA methylation microarrays. *Bioinformatics*. May 15 2014;30(10):1363-9. doi:10.1093/bioinformatics/btu049
